# Impact of COVID-19 Pandemic on Colonoscopy Wait Times by Procedure Indication in Quebec

**DOI:** 10.1101/2024.03.20.24304332

**Authors:** Melina Thibault, Alan Barkun, Myriam Martel, W. Alton Russell

**Affiliations:** Department of Epidemiology, Biostatistics, and Occupational Health, McGill University, Montreal, QC, Canada; Gastroenterology, McGill University, The Montreal General Hospital, GI Division, Montreal, QC, Canada

## Abstract

**Background:** Patients are referred for colonoscopy for symptom assessment, screening, and surveillance. Public health measures to mitigate the spread of the COVID-19 pandemic disrupted services and increased patient delays for colonoscopy services in Quebec. The differential impact of these interruptions by colonoscopy indication is largely unknown.

**Methods:** Using 2018–2022 retrospective clinical data from two high-volume Montreal endoscopy centres and provincial administrative data, we characterized changes in colonoscopy wait times and the proportion of waitlisted patients who were delayed (wait time exceeded provincial guidelines) by procedure indication and demographics. We used regression to examine patient characteristics associated with delayed procedures during pre– and intra-COVID-19 periods. We used time series analysis to characterize trends in the proportion of waitlisted patients delayed.

**Results:** The COVID-19-related public health measures resulted in record-high delays (median increase in wait times of 34-159% across indications). While older patients experienced longer wait times pre-pandemic, intra-COVID-19 wait times increased disproportionately for patients younger than 50. The proportion of waitlisted patients delayed peaked mid-2020 (56.9% for screening; 56.0% for symptom assessment patients). By early 2022, the proportion delayed had fallen to 37.3% for screening patients but remained at 53.8% for symptom assessment patients.

**Conclusions:** In Quebec, intra-COVID-19 colonoscopy delays disproportionately impacted symptom assessment procedures and younger patients. Additional capacity or improved triaging may be needed to address persistent delays. Understanding the effects of the pandemic on colonoscopy services can help inform strategies to mitigate harms from on-going delays in Quebec.

## Introduction

In Quebec, COVID-19 Public Health Emergency measures caused a sharp, temporary reduction in colonoscopy capacities, increasing wait times. Delays in elective colonoscopies (e.g. for cancer screening) can negatively impact patient health outcomes and have costly downstream resource demands.^1,2^ Colonoscopies are vital to the management and diagnosis of digestive pathologies, notably, colorectal cancer (CRC). CRC is the most prevalent non-reproductive neoplasm in Canada.^3,4^ Modelers estimated that pandemic-related delays in colorectal cancer (CRC) detection and treatment during 2020-21 will lead to 60,000 – 70,000 excess life years lost in the next 10 years in Canada.^53/20/2024^ ^7:52:00^ ^AM^

Quebec’s standardized colonoscopy triage sheet (CTS) uses 21 indications to triage colonoscopy referrals into five tiers with associated maximum medically acceptable delays **[Figure S1; Table S1-S2].**^6^ Referral indications fall under three categories: *screening* for CRC, *surveillance* to detect recurrent pre-cancers or disease, and *symptom assessment* for patients with suspected disease.^6,7^ The impact of restricted access to colonoscopy during the COVID-19 pandemic across procedure indications and patient demographic groups has not been reported. In this study, we analyzed patient demographic and clinical characteristics associated with delayed colonoscopy and characterized the evolution of colonoscopy delays by procedure indication throughout the pandemic.

## Methods

Using data from two high-volume endoscopy centres in Montreal, we conducted a retrospective cohort study to analyze wait times and delays in colonoscopies. We used descriptive analyses to profile patient characteristics at time of referral, regression to identify associations between clinical and demographic factors and delays, and time series analysis to examine trends in the proportion of the colonoscopy waitlist that was delayed during the pandemic. Delay analyses were based on the guidelines for maximum delay indicated on the CTS for screening and symptom assessment indications. Delay analyses excluded surveillance indications, for which the CTS contains no maximum delay **[Figure S1]**.

### Study Population and Data Pre-processing

All patients aged 18 or older who underwent a colonoscopy between August 29, 2018, and August 11, 2022, at two hospital-based Montreal endoscopy centres were eligible for inclusion. We included patients’ first colonoscopy with a completed referral sheet. We excluded patients rescheduled due to inadequate bowel preparation or with postal codes that were missing, ambiguous, or from outside of Quebec, or patients classified as priority one on their CTS.

### Data Collection

Patient data were extracted from electronic medical records (sociodemographic characteristics) the CTS (indication, referral date) and pathology database (histological findings). We labelled colonoscopies prior to March 15, 2020, as “pre-COVID-19” and procedures after April 15, 2020, as “intra-COVID-19” **[Tables S1-S2]**. We grouped colonoscopies by indication as: “screening” (e.g. Faecal Immunochemical Test (FIT) positive, referral for family history), “surveillance” (e.g. follow-up colonoscopy), or “symptom assessment” (e.g. gastrointestinal distress) **[Figure S1; Tables S1]**. If multiple indications were selected, we used the one with the highest priority. We labelled patients as delayed once their time on the waitlist exceeded the maximum recommended delay per the CTS.

### Descriptive Analysis

We performed univariate logistic regression, ANOVA (analysis of variance), and chi-squared tests to examine covariates (age, sex, colonoscopy category, body mass index [BMI], neighbourhood-level Material Deprivation Centile (MDC) and Social Deprivation Centile (SDC), and urban vs. rural home address) **[linkage described in Supplement II]**.^8–10^ We conducted two-sided independent-samples t-test to compare pre– and intra-COVID-19 patients wait times (from CTS referral to procedure date) across procedure indications.

### Regression Analyses

We conducted univariable (“unadjusted”) and multivariable (“adjusted”) regressions to compare associations between patient characteristics and delayed procedures by procedure indication [**Supplement III]**.

We used logistic regression to analyse characteristics associated with delayed procedure. Included covariates were COVID-19 period (pre– or intra–), sex, age, MDC, SDC, colonoscopy type, colonoscopy category, and interaction terms.^11^ To investigate the COVID-19 impact on the ***degree*** of delay, we generated “normalized wait times,” defined as the ratio of a patients’ actual wait time to targeted wait time based on their CTS **[Equation 1]**.

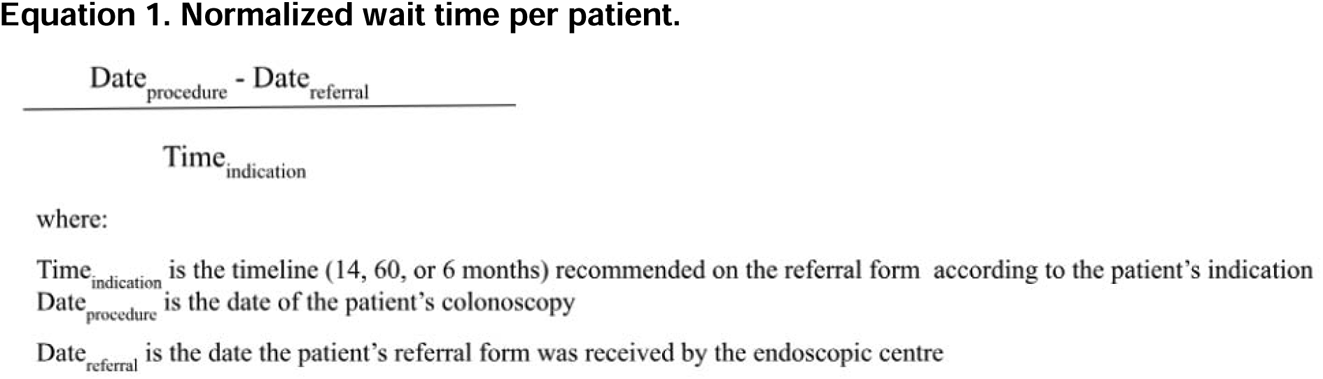

Normalized wait times are ≤1 if patients were not delayed and >1 if they were. We used a generalized linear model with a link function selected using diagnostic tests, considering normal, exponential, logistic, beta, lognormal, and Weibull **[Figure S4-S5]**.^12,13^

We selected regression coefficients using a stepwise selection algorithm with sequential replacement.^14^ We used ANOVA testing on nested models to confirm appropriate covariates. We selected the best-fitting model based on AIC criterion, favouring parsimonious models. Sensitivity analyses and sub-analyses are described in **[Supplement III].**

### Time Series Analysis

We developed seasonal autoregressive integrated moving average models (ARIMA) to describe the proportion of the colonoscopy waitlist delayed for symptom and screening patients over time, accounting for seasonal trends at the weekly, monthly, and yearly level.^15^ We further decomposed our time series data using a robust Multiple Seasonal-Trend decomposition using Loess (MSTL) model,^16–19^ accounting for weekly, monthly, and/or yearly seasonal cycles and aberrant behaviour caused by COVID-19 measures **[Supplement IV]**.

We conducted a counterfactual analysis to estimate the impact of COVID-19 on delayed patients by colonoscopy category. We fit Seasonal Autoregressive Integrated Moving Average with Exogenous Variables (SARIMAX) models using March 13, 2020, as a changepoint for symptom colonoscopies and March 19^th^, 2020, for screening colonoscopies. Changepoints were selected from dates shortly before or after the declaration of the public health emergency in Quebec (March 13th, 2020) using AIC and BIC criterion from candidate SARIMAX models. These models assumed a permanent rise in the proportion of delayed colonoscopies (step change) and a change in slope (ramp). The ramp term was calculated using the mean percent difference in the monthly province-wide dataset after March 13th, 2022. We plotted the counterfactual against observed values for 80 days (e.g. a forecast of how the waitlist would have evolved without the interruption of emergency COVID-19 public health measures, estimated from the pre-changepoint data using SARIMAX modelling).

Full model selection and diagnostic procedures are outlined in **[Supplement IV].** All analyses used R software version 4.1.2. This study was approved by the McGill University Health Centre Research Ethics Board.

## Results

After excluding 20 priority 1 patients, 33 patients from outside Quebec, and 41 with missing or ambiguous postal codes, the dataset included 7,438 pre-COVID colonoscopy procedures and 7,122 intra-COVID procedures. **[Figure 1]**. Average age was higher for surveillance patients compared to symptom or screening, and average age increased among intra-COVID-19 procedures across all three colonoscopy categories **[Table S3-S5]**.

**Figure 1.**
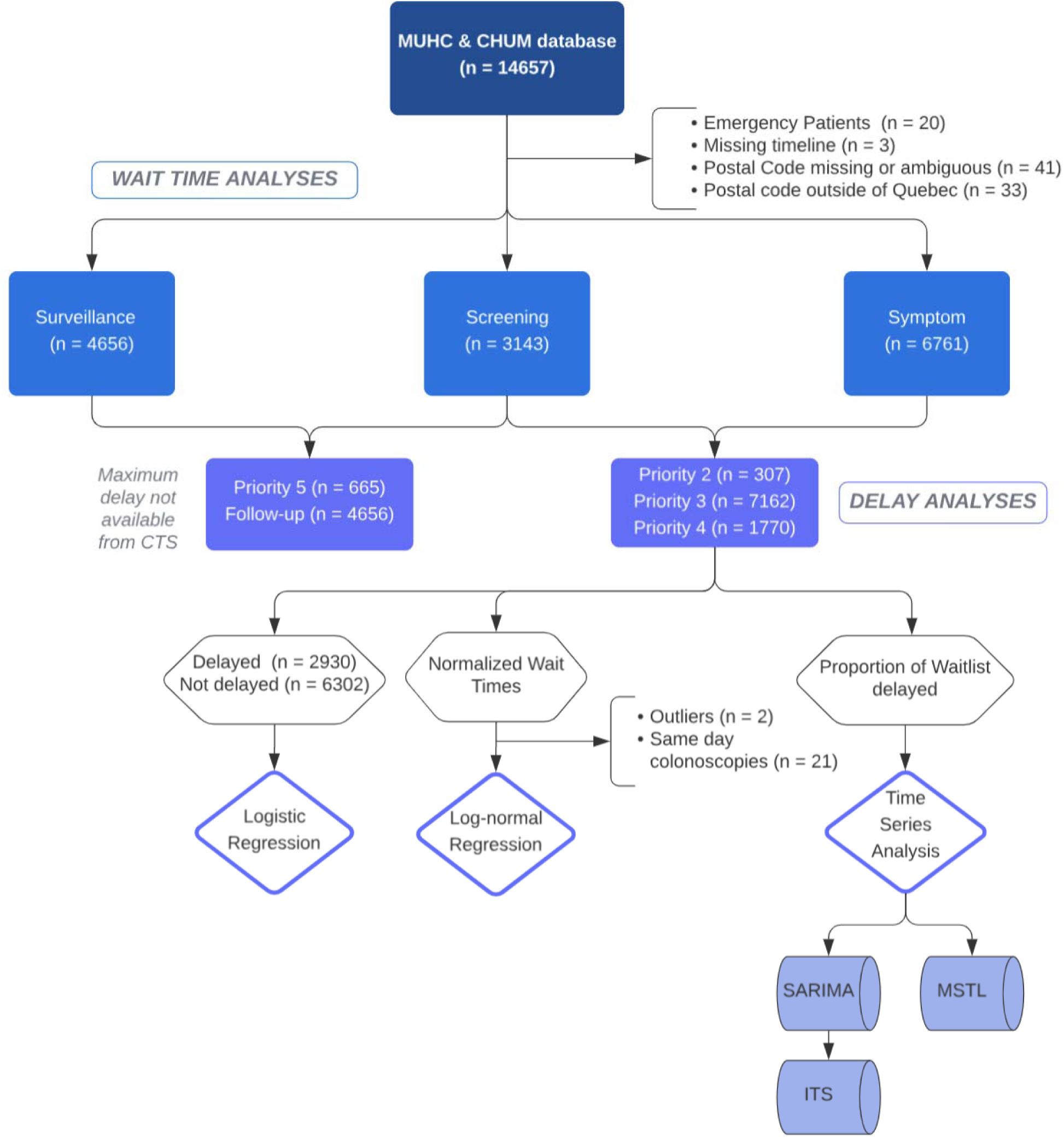
Workflow and Patient Count Diagram. Describes the data management and various analyses conducted during this project as well as the distribution of data based on how different data points were excluded following study inclusion criteria.

On average, patients undergoing colonoscopies for symptom assessment were from more socially and materially deprived neighbourhoods compared with those referred for screening and surveillance indications **[Table 1]**. Neighbourhood deprivation was largely consistent between pre– and intra-COVID periods, except for a small decrease in social deprivation among screening patients **[Table S3-S5]**.

**Table 1.**
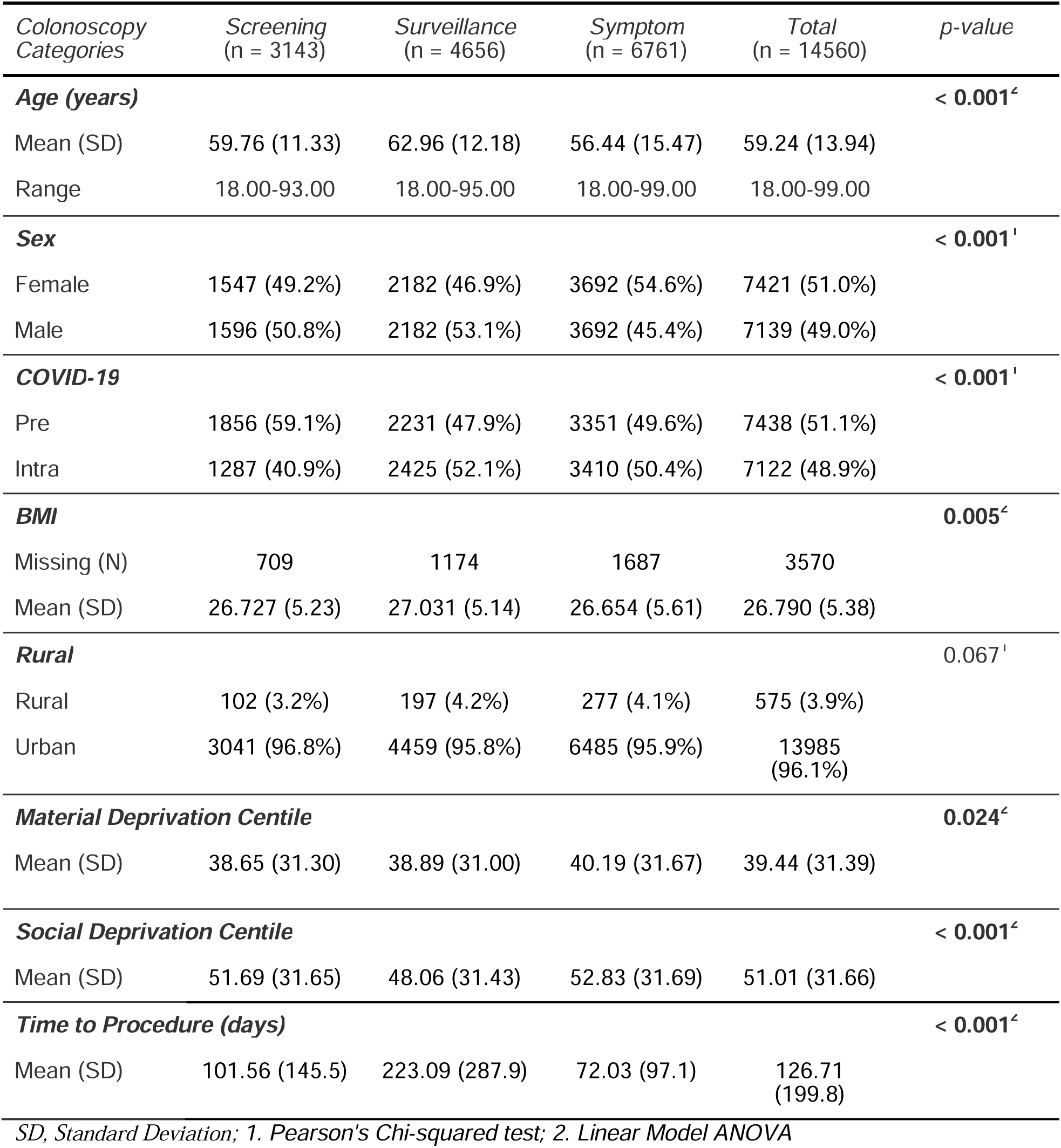
Summary of Socio-Demographic Variables By Colonoscopy Category. Covariates identified from the referral form and linkage performed are investigated per colonoscopy category.

Wait times were longer in the intra-COVID-19 period for all age groups, sexes, and indications apart from male screening patients aged >74 **[Figure 2.b; Figure 2.c]**. Median wait time increased from 46 to 62 days (34% increase) for screening colonoscopies; from 28 to 67 days (139% increase) for symptom assessment; and from 74 days to 192 (159% increase) for surveillance **[Figure 2.a]**. Patients <50 years old experienced the largest increase in proportion of procedures delayed **[Figure 2.b]**. The proportion of delayed procedures increased among screening [increase of 15.2%; CI 14.5 to 16.0; p < 0.001] and symptom assessment indications [increase of 17.2%; CI 16.6, 17.7; p < 0.001]. Detection of clinically significant legions was higher for rural patients (45.2% vs 39.0%, p < 0.05) **[Table S7; Figure S2]** and was higher intra-COVID-19 for symptom assessment patients (28.7% to 34.6%; p < 0.001) **[Table 2].**

**Figure 2.**
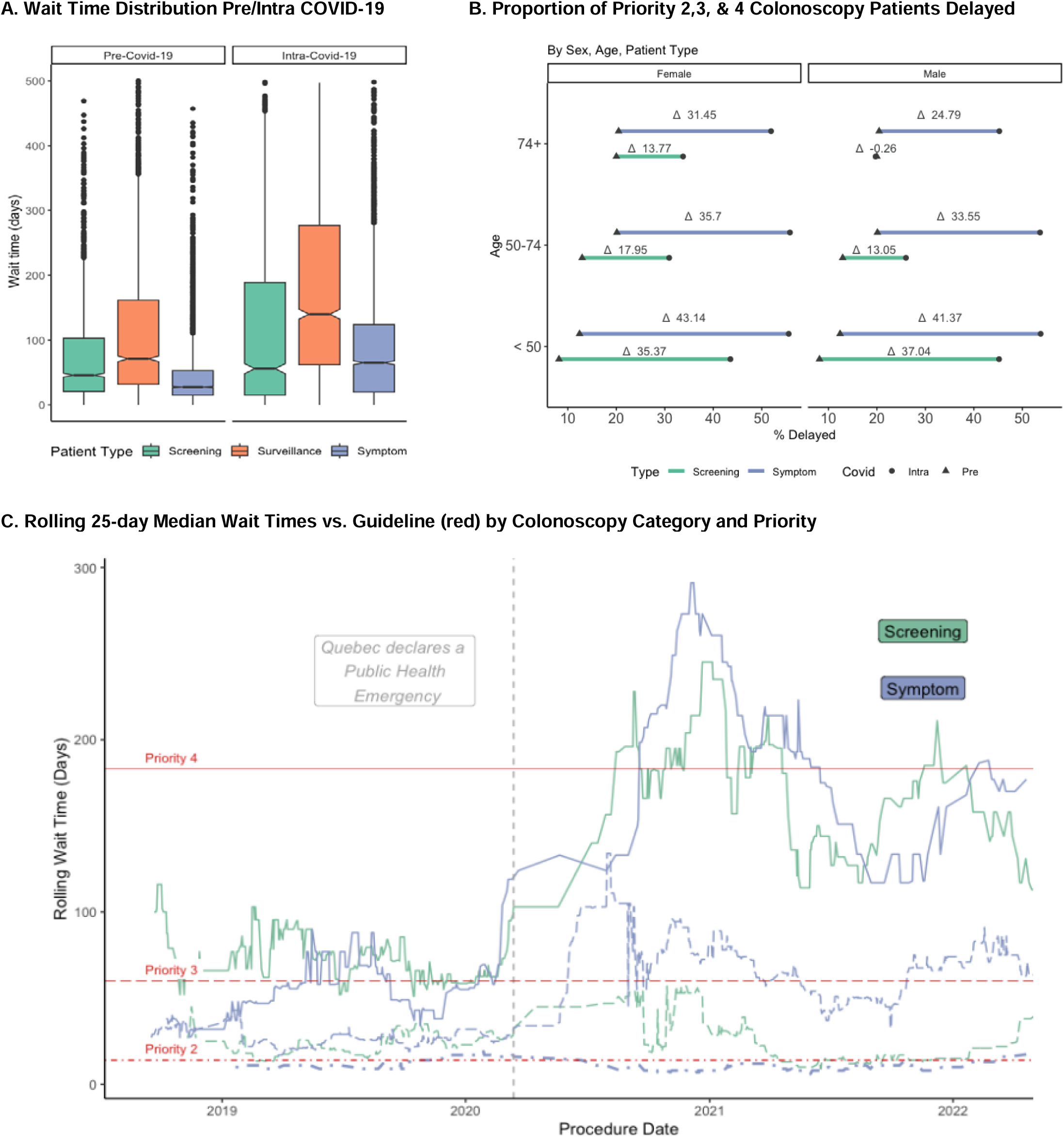
Patient Wait Times and Delays. **A. Wait Time Distribution in Days Pre/Post Covid Across Patient Populations.** The y-axis of the figure is limited to 500 days to highlight the differences in patient wait time distribution. There were 70 pre-COVID-19 patients and 513 intra-COVID-19 patients with wait times exceeding 500 days. Notches display the 95% CI around the median patient wait time. Considering notches of boxes do not overlap, this provides evidence of a statistically significant difference between the medians both across patient subgroups and across pre/intra COVID-19 periods. **B. Proportion of Priority 2, 3, & 4, Patients Delayed Across Screening and Symptom Patients by Sex and Age**. The proportion delayed of all patients stratified by COVID-19 period, age, and colonoscopy category is shown. The percent difference (delta) in patients delayed is noted]. **C. 25-day Rolling Median Wait Times for Priority 2, 3, & 4 Patients by Colonoscopy Category.** Red lines mark the recommended wait time on the CTS per priority group. Pre– and Intra-Covid-19 median wait times in days per screening and symptom assessment patient groups are shown with priority subtypes. From top to bottom: Screening and Symptom Priority 4 (solid line), Symptom and Screening Priority 3 (dashed line), Symptom Priority 2 (dot-dash line). We see Symptom Priority 3 patients have increased median wait times compared to Screening Priority 3 patients.

**Table 2.** Summary of Colonoscopy Findings By Colonoscopy Category. A patient was deemed to have clinically significant findings if their procedure resulted in the identification of lesions, including acute colitis, polyps [any type], ileocolitis, diverticulosis, haemorrhoids (if this was the presumed source of bleeding), Crohn’s, ulcerative colitis, infectious or pseudomembranous colitis, radiation colitis, ischemic colitis, solitary ulcer, vascular lesions, strictures, microscopique colitis, collagenous colitis, adenoma, and/or adenocarcinoma.

| *Colonoscopy Categories* | *Screening*<br>*(n = 3143)* | *Surveillance*<br>*(n = 4656)* | *Symptom*<br>*(n = 6761)* | *Total*<br>*(n = 14560)* | *p-value* |
| --- | --- | --- | --- | --- | --- |
| **Cancer** |  |  |  |  | **< 0.001<sup>1</sup>** |
| Prevalence | 43 (1.4%) | 13 (0.3%) | 70 (1.0%) | 126 (0.9%) |  |
| **Clinically Significant Lesions** |  |  |  |  | **< 0.001<sup>1</sup>** |
| Prevalence | 1371 (43.6%) | 2205 (47.4%) | 2142 (31.7%) | 5718 (39.3%) |  |

*SD, Standard Deviation; 1. Pearson's Chi-squared test*

### Predictors of Delayed Procedure

Among symptom assessment and screening indications (n=9237), 784 pre-COVID-19 (16.3%) and 2142 intra-COVID-19 procedures (48.6%) were delayed.

In the adjusted model, intra-COVID-19 procedures had higher odds of delay [OR 4.73, 95% CI 4.30, 5.22, p < 0.001] and female patients experienced on average 11% greater odds of having a delayed procedure than males [OR: 1.11 95% CI (1.01, 1.22) p = 0.042]. While older patients experienced greater delays on average. Estimating the interaction between age and the COVID-19 period found that increased age was protective against delay during the intra-COVID-19 period: a 10-year increase in age yielded a 23% reduction in odds of having a delayed intra-COVID-19 procedure in the fully adjusted model [OR: 0.77 95% CI (0.72, 0.82) p < 0.001] **[Table 3]**. Sub-analyses revealed that patients with positive FIT were on average older and experienced less delay compared to screening patients without positive FIT **[Figure S3]**.

**Table 3.** Logistic Regression Models for Delayed Procedures. A univariable (“unadjusted”) regression is noted, with no adjustment per covariate. Two multivariable (“adjusted”) regression models are displayed, adjusting for covariates: COVID-19 period (pre– or intra-), sex, age, Material Deprivation Centile (converted to dentile), Social Deprivation Centile (converted to dentile), colonoscopy type, and interaction terms. The fully adjusted logistic regression model includes an interaction term between COVID-19 period and age.

|  |  | Model without interaction term |  |  | Model with interaction term |  |
| --- | --- | --- | --- | --- | --- | --- |
| Covariate | n | Unadjusted OR<sup>1</sup> | Adjusted OR <sup>1</sup><br>(95% CI) | p-value | Adjusted OR <sup>1</sup><br>(95% CI) | p-value |
| COVID-19 |  |  |  |  |  |  |
| Pre | 4 816 | Ref. | Ref. | - | Ref. | – |
| Intra | 4 421 | 4.86 (4.41-5.35) | 4.74 (4.30-5.23) | **<0.001** | 21.81 (14.55-32.84) | **< 0.001** |
| Colonoscopy Categories |  |  |  |  |  |  |
| Screening | 2 478 | Ref. | Ref. | - | Ref. | – |
| Symptom | 6 759 | 2.29 (2.05-2.56) | 2.18 (1.94-2.46) | **<0.001** | 2.18 (1.94-2.45) | **< 0.001** |
| Sex |  |  |  |  |  |  |
| Male | 4 311 | Ref. | Ref. | - | Ref. | – |
| Female | 4 926 | 1.14 (1.05-1.25) | 1.11 (1.01-1.22) | **0.037** | 1.10 (1.00-1.21) | **0.045** |
| Age <sup>2</sup> |  |  |  |  |  |  |
| Per decade | - | 1.04 (1.01-1.07) | 1.04 (1.01-1.07) | **0.016** | 1.23 (1.16, 1.30) | **< 0.001** |
| Deprivation Dentile <sup>2</sup> |  |  |  |  |  |  |
| Material | - | 0.99 (0.98-1.01) | 0.99 (0.97, 1.00) | 0.147 | 0.99 (0.97, 1.00) | 0.124 |
| Social | - | 1.00 (0.99-1.02) | 1.01 (1.00, 1.03) | 0.187 | 1.01 (0.99, 1.02) | 0.219 |
| Covid-19 * Age <sup>2</sup> |  |  |  |  |  |  |
| Pre * Age | - | - | - | - | Ref. | – |
| Intra * Age | - | - | - | - | 0.77 (0.72, 0.82) | **< 0.001** |

OR Odds ratio; CI, Confidence Interval; Ref. referent category
<sup>1</sup> Exponentiated regression coefficient; <sup>2</sup> Covariates were rescaled by 10

### Predictors of Normalized Wait Times

Pre-COVID-19, most patients were seen on time (normalized wait time ≤1). In the intra-COVID period, normalized wait times increased to 0.55 (+0.14) for screening patients and to 1.1 (+0.65) for symptom patients **[Table S9].**^20–22^

From our log linear regression model, normalized wait times increased by 0.76 [Adjusted estimate 1.76 95% CI (1.68, 1.84) p < 0.001] for patients with intra-COVID-19 times **[Table 4]**. Age was strongly associated with increases in normalized wait times, with an 11% increase per 10-year increase in age [adjusted estimate 1.11 95% CI (1.08, 1.13) p < 0.001]. Additionally, there was a significant crossover interaction between COVID-19 period (pre– or intra-) and age [0.89 (95% CI 0.86, 0.91) p < 0.001. **[Figure 3]**. This interaction indicates a reversal of the effect of age on wait times across strata of the COVID-19 period.

**Figure 3:**
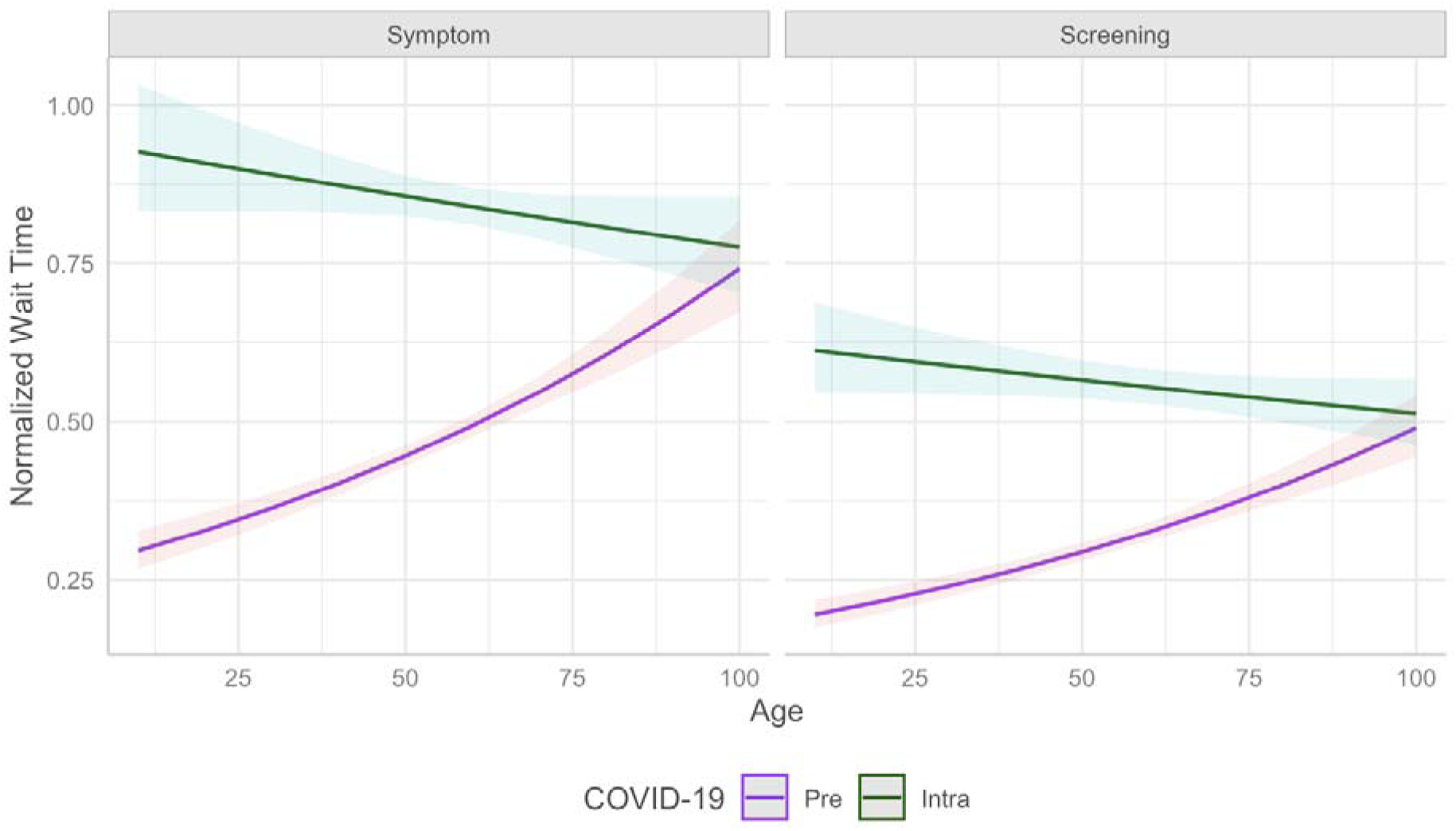
Marginal Effects of Age and COVID-19 in Log-Normal Regression. Older age was associated with longer wait times in the pre-COVID period, but this trend reversed in the intra-COVID period.

**Figure 4.**
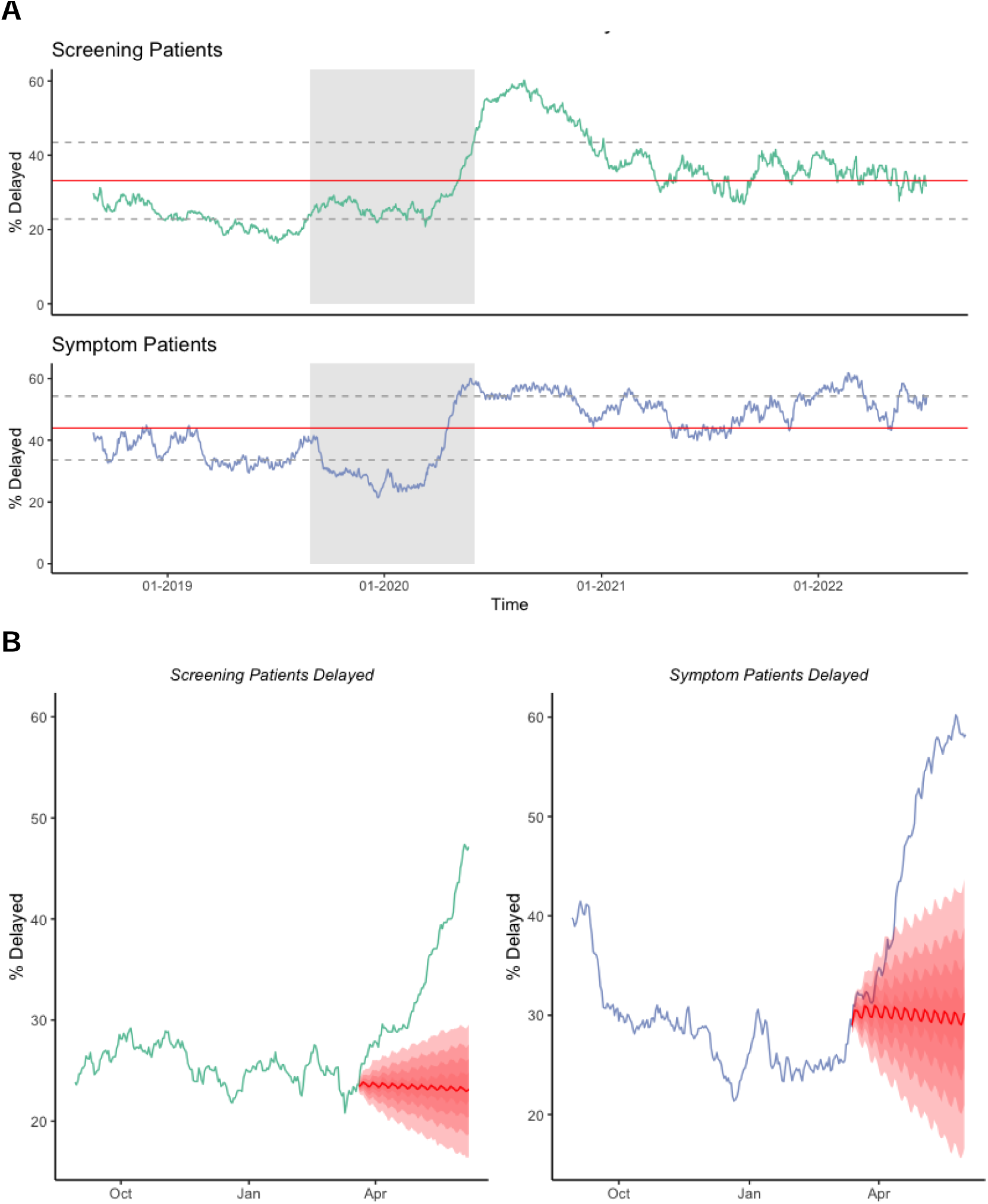
SARIMAX Models. A. Raw Time Series of Proportion of Waitlist Volume Delayed Per Patient Population. The red line is the mean of the proportion delayed over time and the dotted lines indicate the standard deviation. A steep increase is noted in 2020 for both patient populations following the beginning of the COVID-19 public health emergency in Quebec. The shaded regions highlight the time window noted in section B.] **B. Counterfactual Plots (2019-2020): Observed vs Predicted:** Data is plotted for August 2019 till June 2020. The counterfactual is in red against the observed data per patient population waitlists for 80 days following the change point. It shows the average predicted trend in this counterfactual with associated prediction intervals (20%, 40%, 60%, and 80%). Weekly seasonality is captured in the predicted line.

**Table 4.** Log-Linear Regression Models for Normalized Wait Times. A univariable (“unadjusted”) regression is noted, with no adjustment per covariate. Two multivariable (“adjusted”) regression models are displayed, adjusting for covariates: COVID-19 period (pre– or intra-), sex, age, MDC dentile, SDC dentile, colonoscopy type, and interaction terms. The fully adjusted log-linear regression model includes an interaction term between COVID-19 period and age.

|  |  | Model without interaction term |  |  | Model with interaction term |  |
| --- | --- | --- | --- | --- | --- | --- |
| Covariate | n | Unadjusted OR<sup>1</sup> | Adjusted Estimate<sup>1</sup><br>(95% CI) | p-value | Adjusted Estimate <sup>1</sup><br>(95% CI) | p-value |
| COVID-19 |  |  |  |  |  |  |
| Pre | 4 793 | Ref. | Ref. | - | Ref. | – |
| Intra | 4 388 | 1.82 (1.74-1.91) | 1.76 (1.68-1.84) | <0.001 | 3.53 (2.96-4.22) | < 0.001 |
| Colonoscopy Category |  |  |  |  |  |  |
| Screening | 2 471 | Ref. | Ref. | - | Ref. | – |
| Symptom | 6 710 | 1.57 (1.49-1.66) | 1.51 (1.44-1.59) | <0.001 | 1.51 (1.44-1.59) | < 0.001 |
| Sex |  |  |  |  |  |  |
| Male | 4 287 | Ref. | Ref. | - | Ref. | – |
| Female | 4 894 | 1.06 (1.01-1.11) | 1.03 (0.99-1.08) | 0.140 | 1.03 (0.99-1.08) | 0.157 |
| Age <sup>2</sup> |  |  |  |  |  |  |
| Per decade | - | 1.04 (1.02-1.06) | 1.04 (1.03-1.06) | <0.001 | 1.11 (1.08, 1.13) | < 0.001 |
| Deprivation Dentile <sup>2</sup> |  |  |  |  |  |  |
| Material | - | 0.99 (0.99-1.00) | 0.99 (0.99-1.00) | 0.018 | 0.99 (0.98, 1.00) | 0.013 |
| Social | - | 1.00 (0.99-1.01) | 1.01 (1.00-1.01) | 0.159 | 1.00 (1.00, 1.01) | 0.178 |
| Covid-19 * Age <sup>2</sup> |  |  |  |  |  |  |
| Pre * Age | - | - | - | - | Ref. | – |
| Intra * Age | - | - | - | - | 0.89 (0.86, 0.91) | < 0.001 |

OR Odds ratio; CI, Confidence Interval; Ref. referent category
<sup>1</sup> Exponentiated regression coefficient
<sup>2</sup> Covariates were rescaled by 10

### Time Series Analysis

For both screening and symptom assessment indications, counterfactual SARIMAX models suggested that without the pandemic, the proportion of colonoscopies delayed would have decreased slightly in the 80 days after the public health emergency was declared (to 30.0% for symptom assessment; to 23.4% for screening). Instead, they increased (to 50.9% for symptom assessment; 39.4% for screening). After adjusting for time-varying weekly and monthly patterns, MSTL analysis uncovered a peak in the proportion of screening procedures delayed in August 2020 (56.9% of waitlist delayed) followed by a decline. The proportion of screening procedures delayed has still not returned to pre-pandemic levels (mean proportion delayed of 22.7% in 2019 compared to mean proportion delayed of 37.3% in the first 6 months of 2022) **[Figure 5]**. For symptom procedures, MSTL analysis showed a slow decline in proportion delayed pre-COVID-19, followed by a sharp increase in 2020 that peaked at 56.0% in May 2020). The proportion delayed did not return to pre-pandemic levels (53.8% in the first 6 months of 2022 vs 33.6% in 2019).

**Figure 5.**
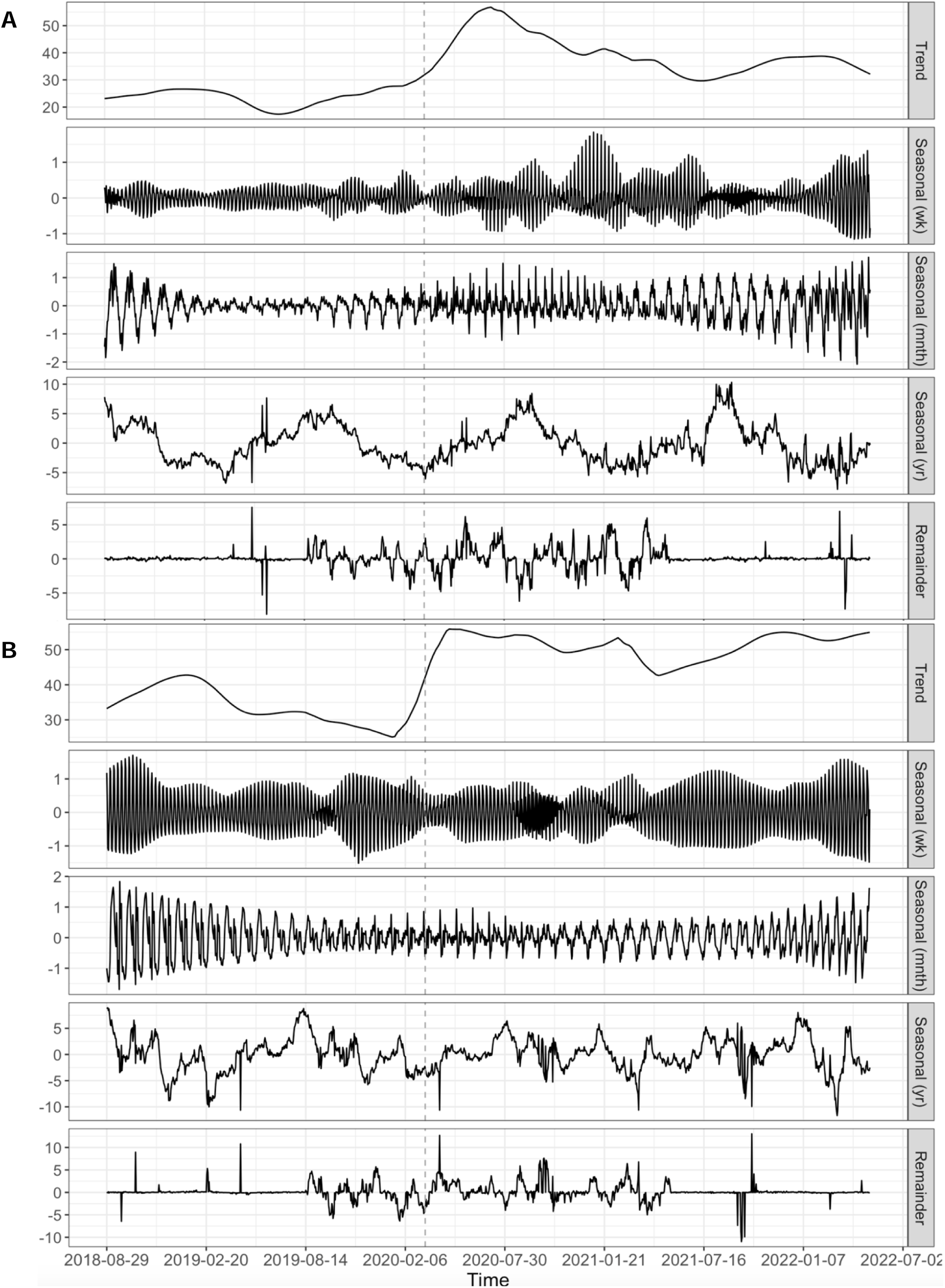
MSTL Models. A. Screening MSTL. The resulting screening MSTL showed increased variance around the beginning of the intra-COVID-19 period. Annual seasonality is noted, with peaks in the late summer, early fall. **B. Symptom MSTL.** The symptom MSTL similarly described increased variance around the start of the intra-COVID-19 period. Annual seasonality is not significant, approaching a white noise pattern.

## Interpretation

In 2020, public health COVID-19 measures in Quebec interrupted routine medical care, including colonoscopies.^20,21^ We found a sharp climb in delayed procedures for screening and symptom patients in 2020. In 2022, the proportion of patients delayed remained higher than pre-pandemic levels, particularly for symptom assessment indications.

Older patients had greater delays in care and treatment pre-COVID-19, but increased delays during the intra-COVID period disproportionately impacted patients under 50. Patients under 50 are not included in CRC screening protocols in Canada, although CRC screening ages have been lowered in the US due to increasing rates of CRC.^23,24^ The disproportionate impact on younger patients could have several causes. Firstly, early pandemic messaging encouraging telehealth alternatives over in-person primary care may have inadvertently led young patients to delay colonoscopies.^25^ Secondly, young people may have developed greater healthcare avoidant behaviours over the pandemic,^26^ however evidence from other countries is mixed.^27^ Lastly, the healthcare system prioritized FIT positive patients^28^, who tend to be older^29^ and carry a heightened risk of CRC findings, and worsened oncological prognoses.^30^

While FIT is considered to have more clinical and analytical sensitivity over other tests in detecting occult blood, a marker for neoplasms,^31^ how the prioritization of FIT patients impacted other colonoscopy-seeking patients has not been explored. FIT positive patients in Quebec may constitute a higher risk population than participants in other screening programs,^32^ as the province has the highest positivity threshold across Canada (175ng/mL),^33^ which may support their prioritization. Our study suggests that with finite colonoscopy resources, wait times for other indicated conditions may have been displaced by this reorganization. Indeed, the degree of delay noted in our analyses supports smaller increases in wait times for FIT patients (e.g. screening patients).

Intra-COVID-19 patients saw increased wait times irrespective of sociodemographic characteristics. Though disparities were small, we found that women had slightly longer wait times compared to men, and patients from more materially deprived neighbourhoods had longer normalized wait times. These are concerning trends, given reports of socio-cultural barriers in CRC screening for women^34^ and the disproportionate burden of CRC among those in Canadian low neighbourhood income quintiles.^3^

Province-wide data indicates continued delays in colonoscopy access, as in several countries.^2,35–37^ As of August 12, 2023, roughly 130,000 patients were on the waitlist for colonoscopies, compared to 59,000 on February 29, 2020.^38^ Chronic understaffing, mandated overtime, and increased rates of burnout across healthcare workers continue to undermine efforts to combat waitlist delays.^39^

Our study had several limitations. Selection into the study was based on patients having a referral form, which is a suggested but not compulsory practice used by >70% of referral centres in Quebec.^40^ Systematic differences between patients referred with and without the form could bias our findings. Patients who were referred but never seen for a colonoscopy were not included, so we likely underestimated the proportion of waitlisted patients delayed. Given the relative urgency of symptom assessment and screening patients, we estimate that patients most impacted by this bias were those referred for surveillance indications, which were not our focus. Furthermore, the lack of a set deadline for Priority 5 and Follow-up colonoscopies, meant an inability to include them in our delay analysis. Future studies could use surveillance patients’ complete endoscopic history to determine whether procedures exceeded wait time guidelines.

This is the first study of its kind in the province. While data was constrained to two endoscopic centres, comparisons with administrative data indicated consistent secular trends across the province **[Figure 6]**. Province-wide digitization of colonoscopy records could enable monitoring of colonoscopy access across patient groups, enabling more granular evaluation of the impact of interventions to mitigate harms from delayed procedures.

**Figure 6.**
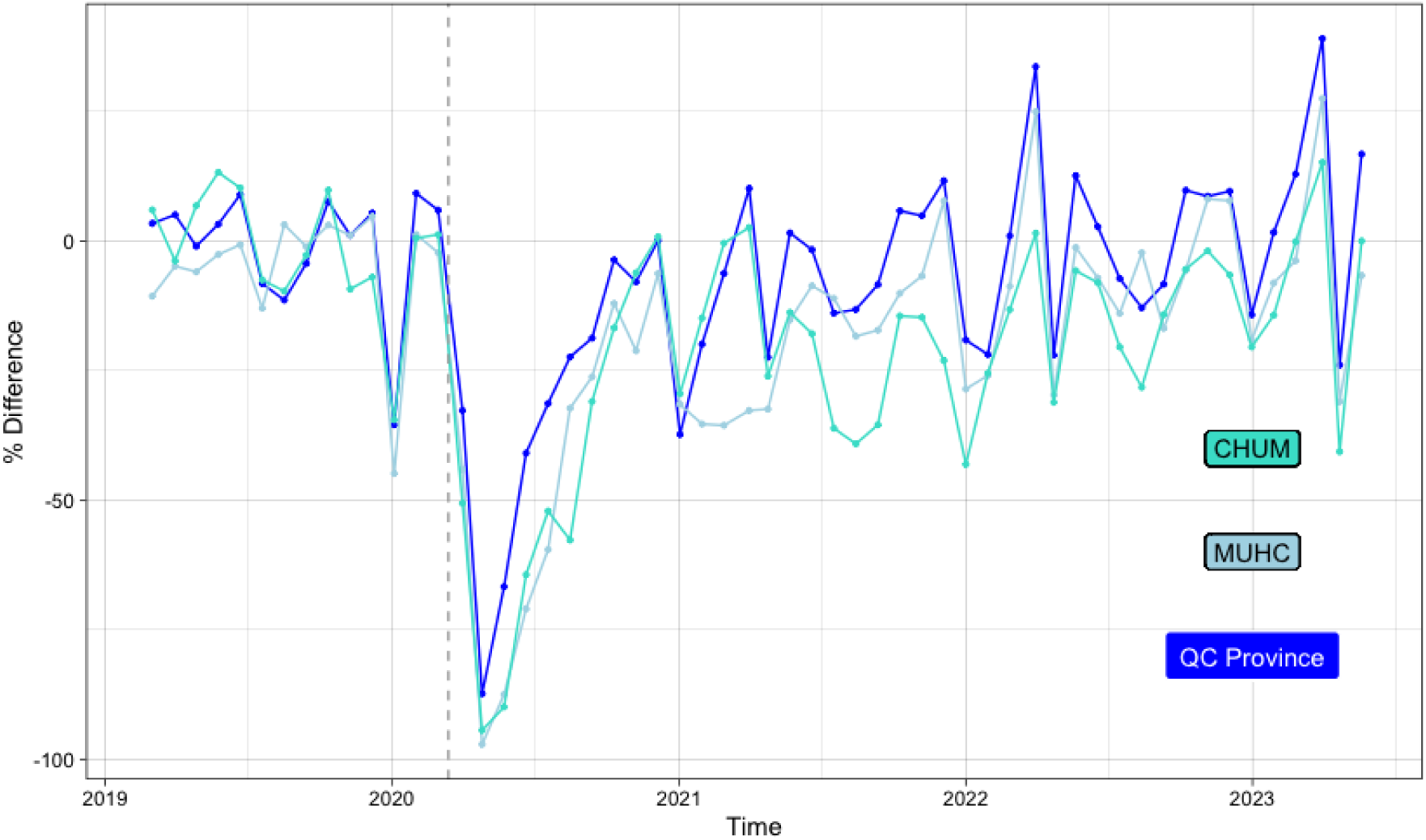
Monthly Colonoscopies in Administrative Provincial Data compared to Endoscopy centres. Endoscopy-providing centres in Quebec, regularly report the number of colonoscopies conducted and FIT administered by fiscal period (13 per year). Reporting is at the discretion of clinics, and some choose to only report symptomatic and screening colonoscopies, excluding surveillance indications. The province-wide data is amalgamated by the provincial ministry and disseminated through a PowerBI dashboard.^38^ Other data reported includes colonoscopy capacity per centre and the burden of CRC disease. Colonoscopy volumes per financial period was compared to the mean number of colonoscopies performed per reporting body in 2018 and compared across provincial and individual centres’ administrative data. There are noted drops in colonoscopy volumes through the holiday period annually (Dec-Jan) and for spring break (Apr-Mar). The sharp decline in colonoscopies following the implementation of COVID-19 public health emergency measures is noted. Similarly, the incline in colonoscopies services as normal working hours resumed is also reflected. We see and effort to increase colonoscopy capacity in 2022-2023.

Our study offers insight into the effects of COVID-19 on colonoscopy wait times, highlighting significant increases in wait times for certain patient groups and factors associated with delays. While acknowledging limitations, our analysis focused on how public health decisions, especially the triaging of FIT positive referrals and prioritization of older age groups, affected patient access to healthcare services. These results underscore the demand for healthcare strategies that could both lessen the impact of future disruptions on colonoscopy services and ensure timely access to care for patients. They also highlight the need for accessible information, potentially through digitized patient records, to inform these decisions.

## Data Availability

Data not available due to ethical restrictions

## Acknowledgements

Thank you to our collaborators, *Ministère de la Santé et des Services sociaux*, especially Oronzo de Benedictis, for their help with the provincial dataset. Similar gratitude is expressed to l’*Institut national d’excellence en santé et en services sociaux*.

