## Supplement Figures for "Impact of COVID-19 Pandemic on Colonoscopy Wait Times by Procedure Indication in Quebec"

#### **Figure S1. Colonoscopy Triage Sheet.** The CTS form used to triage patients in Quebec following referral for a colonoscopy.

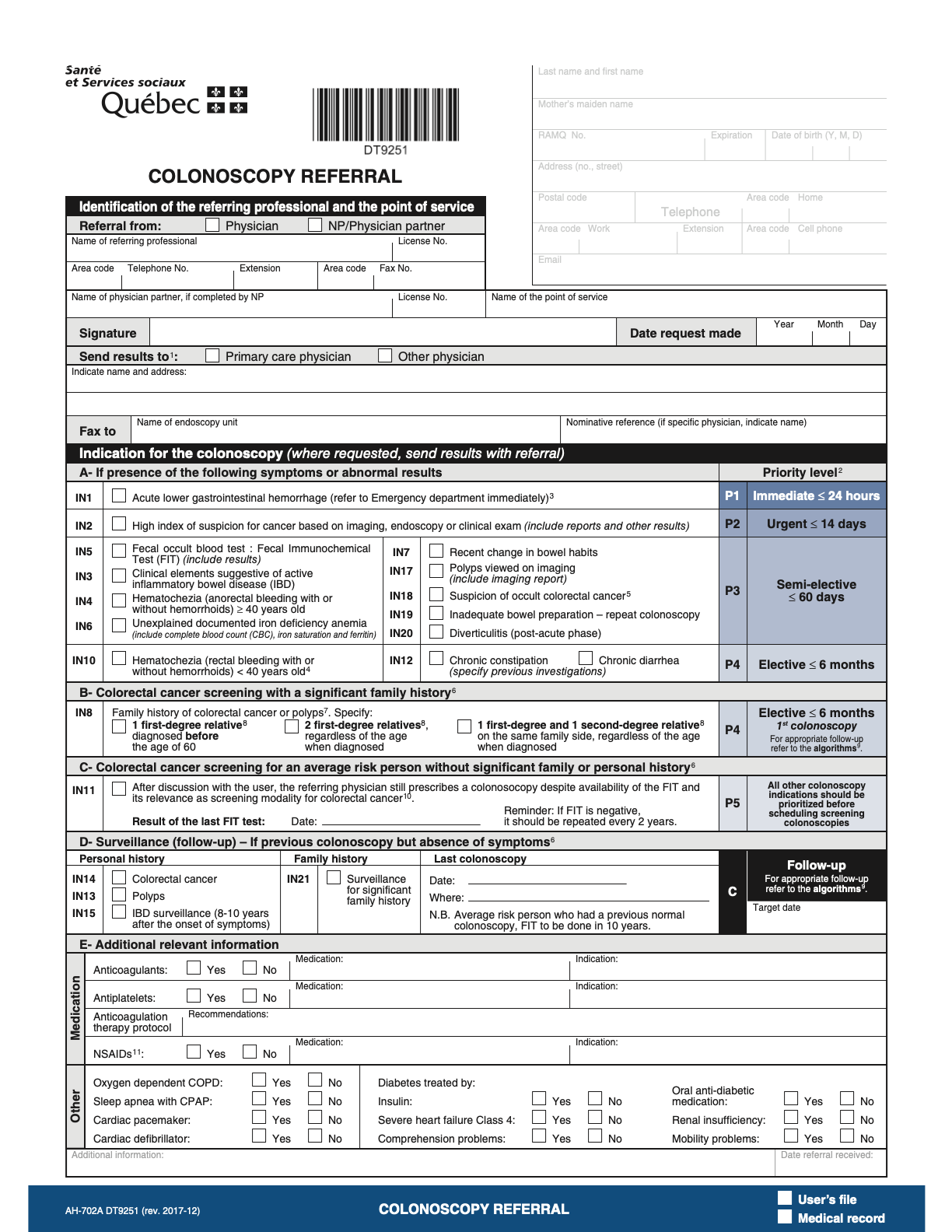

#### **Figure S2. Patient’s Neighbourhood Deprivation Indices Distributions Pre/Intra COVID-19 per Colonoscopy Category**

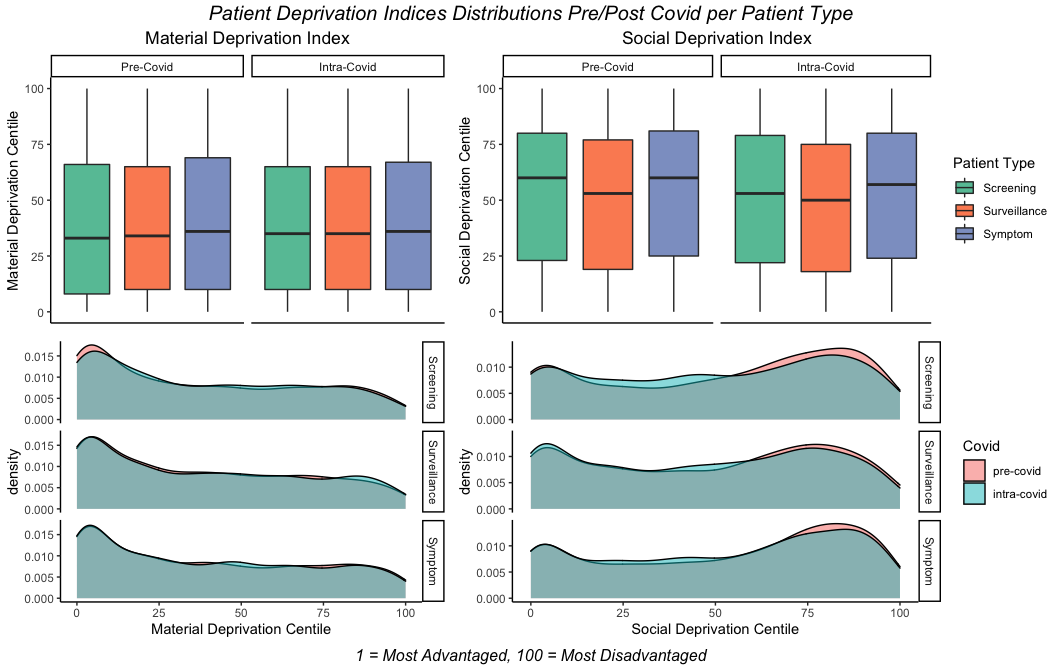

#### **Figure S3. Interaction between Age, FIT and COVID-19**

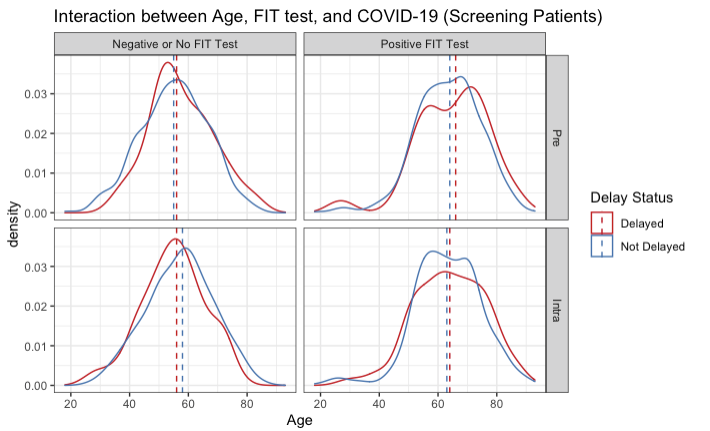

#### **Figure S4. Cullen and Frey Graph**

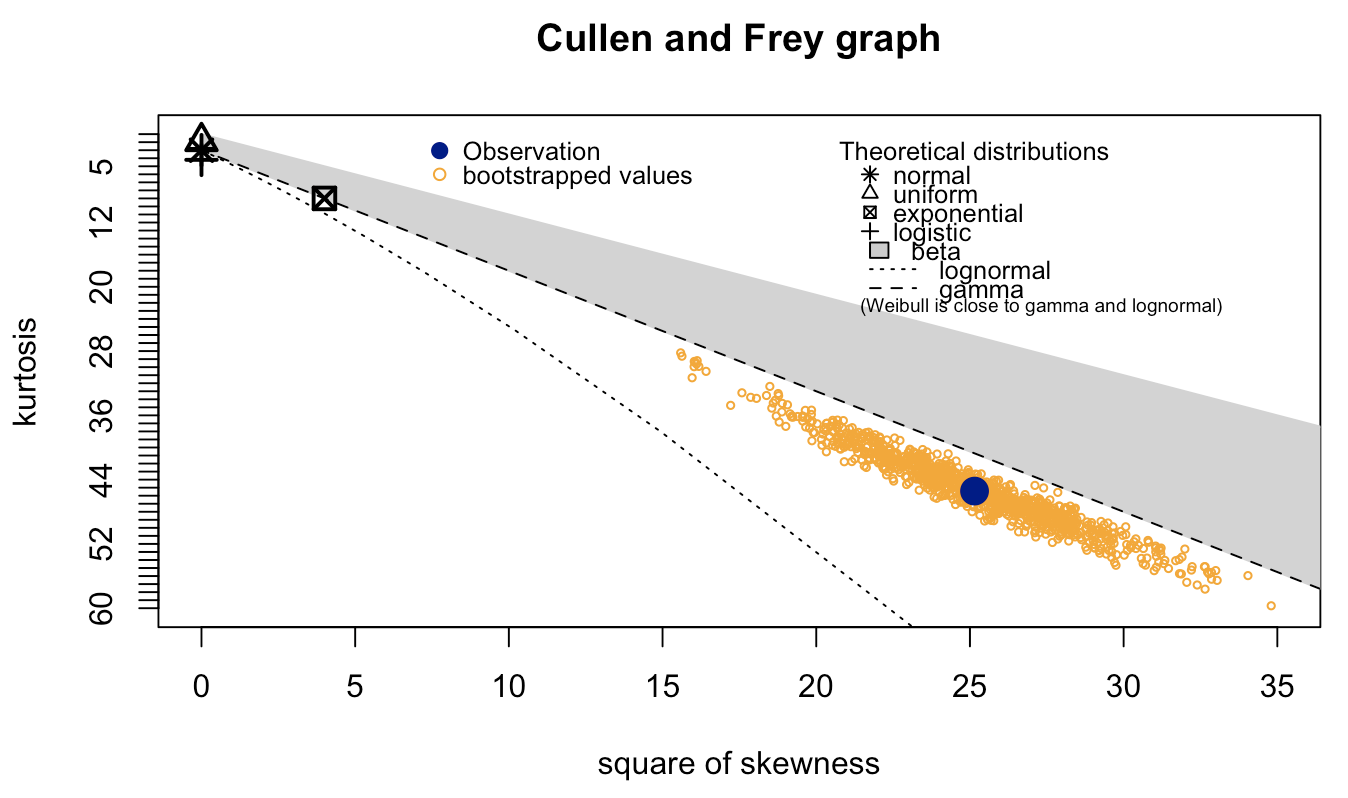

#### **Figure S5. Goodness of Fit Plots**

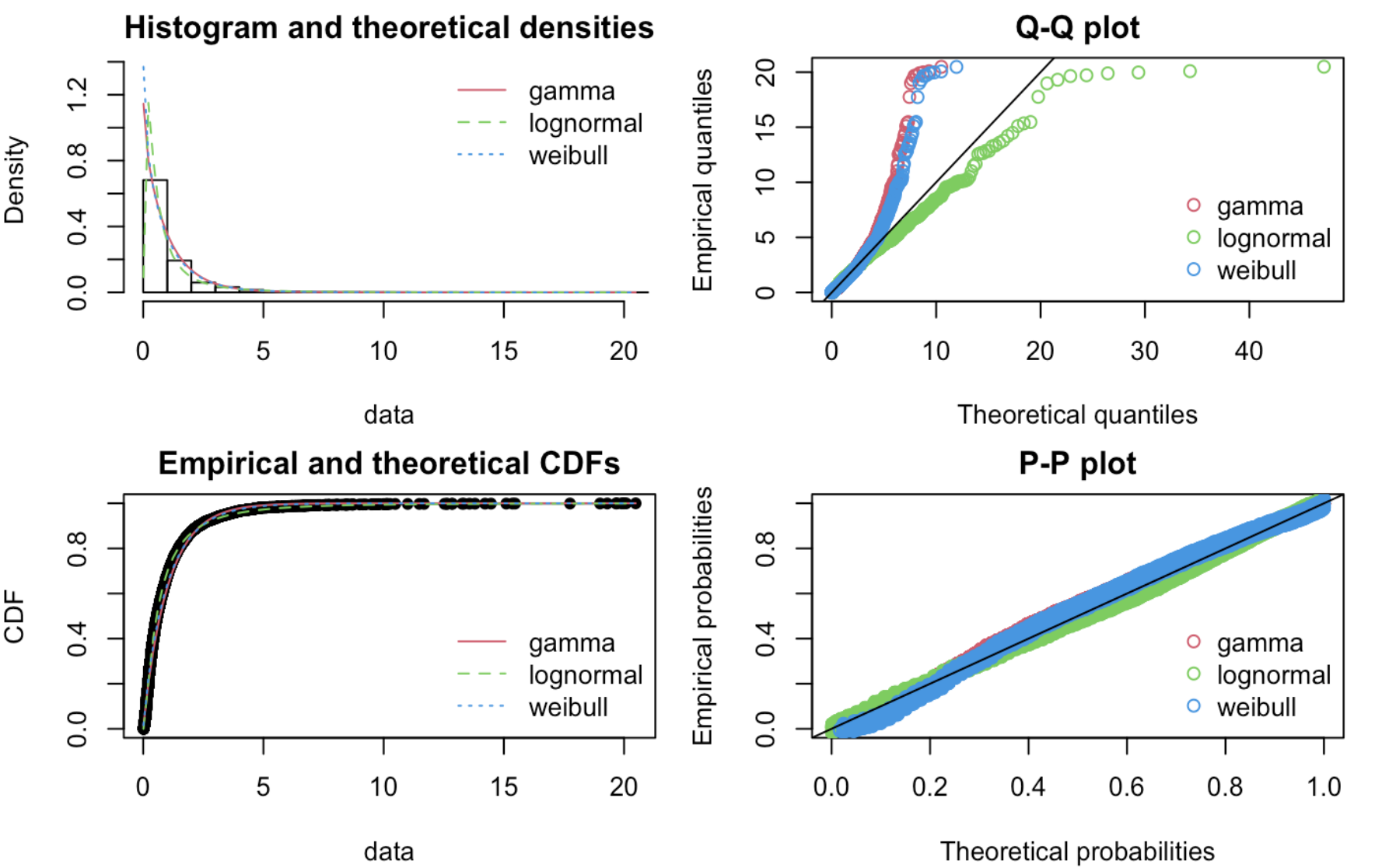

#### Figure S6. Diagnostic Checks for Log Normal Model

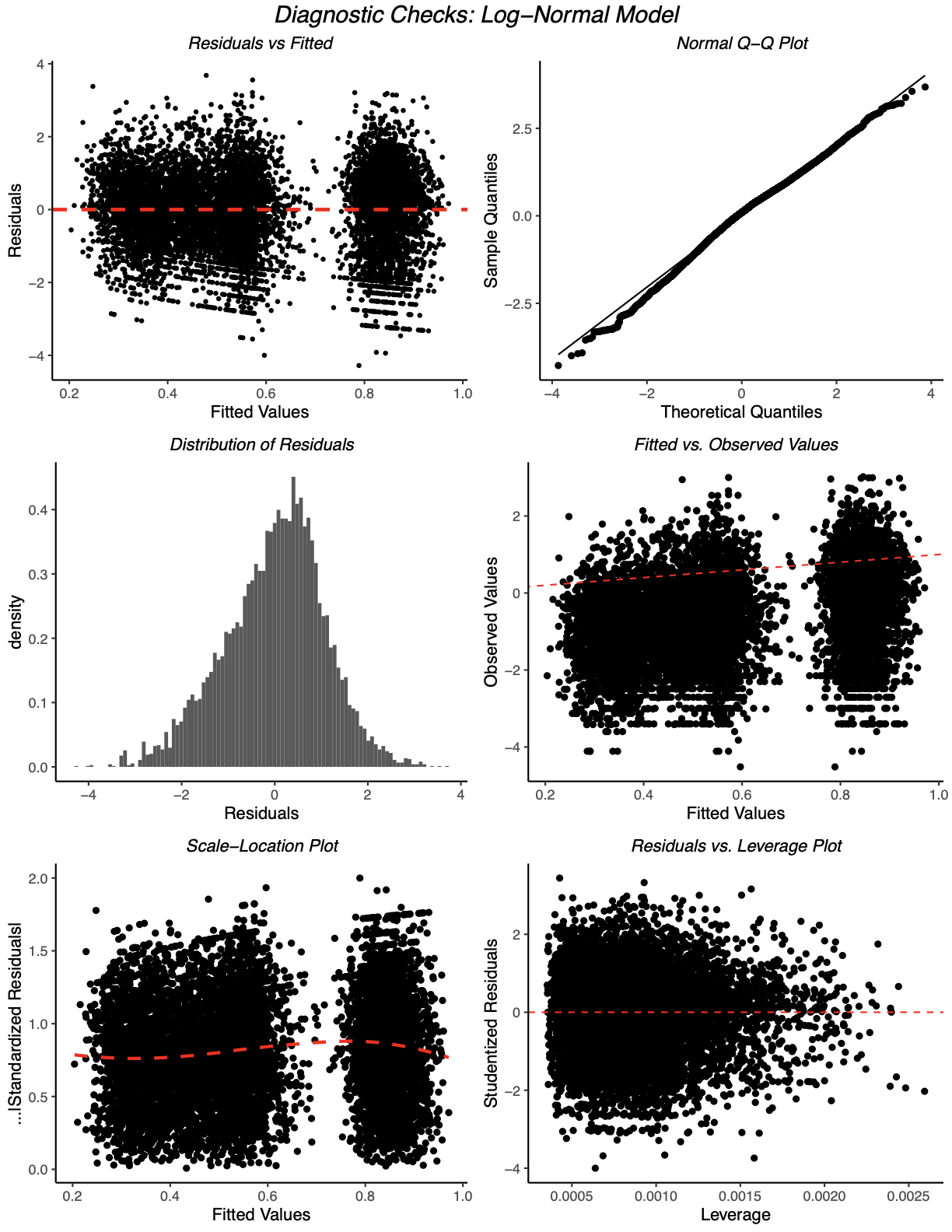

**Figure S7. Distribution of Normalized Wait Times per Colonoscopy Category.**

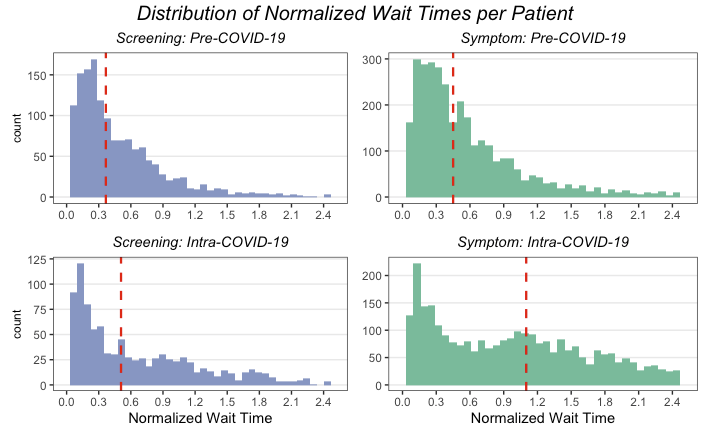

**Figure S8. Residuals By COVID-19 Period**

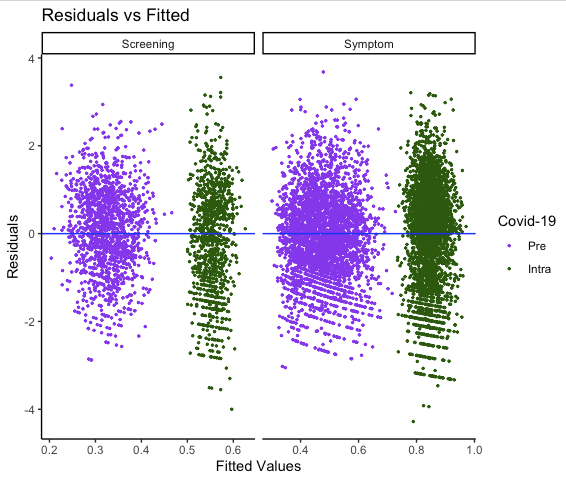

**Figure S9. How proportion of waitlist delayed was calculated.**

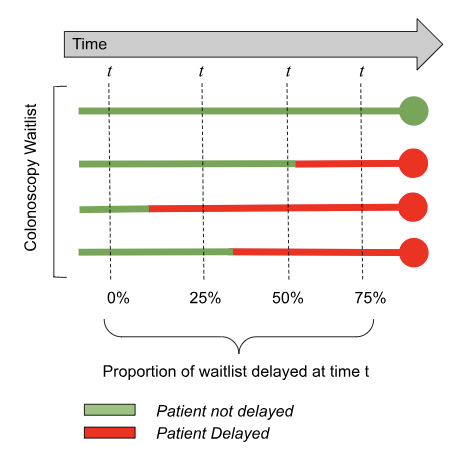

**[Figure S9. How proportion of waitlist delayed was calculated.** Each line represents a patient's trajectory on the colonoscopy waitlist.]

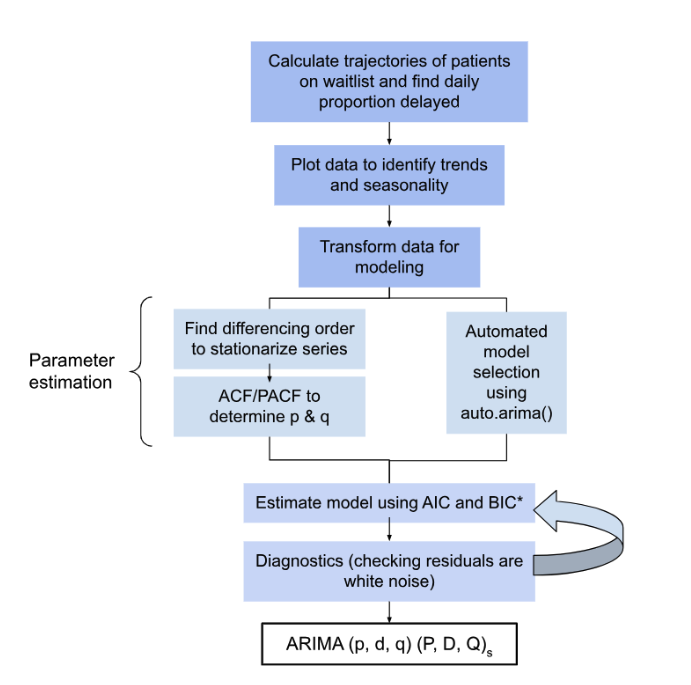

**Figure S10. Workflow for ARIMA modelling.** Diagram for model selection adapted from Hyndman and Athanasopoulos**. ***Considering the aim of this paper was not prediction, the goal was to conserve interpretability by minimizing model fit statistics and retaining the smallest AR and MA terms.

**Figure S11. ACF/PACF Plots**

A. **ACF Plots: Autocorrelation Functions.**

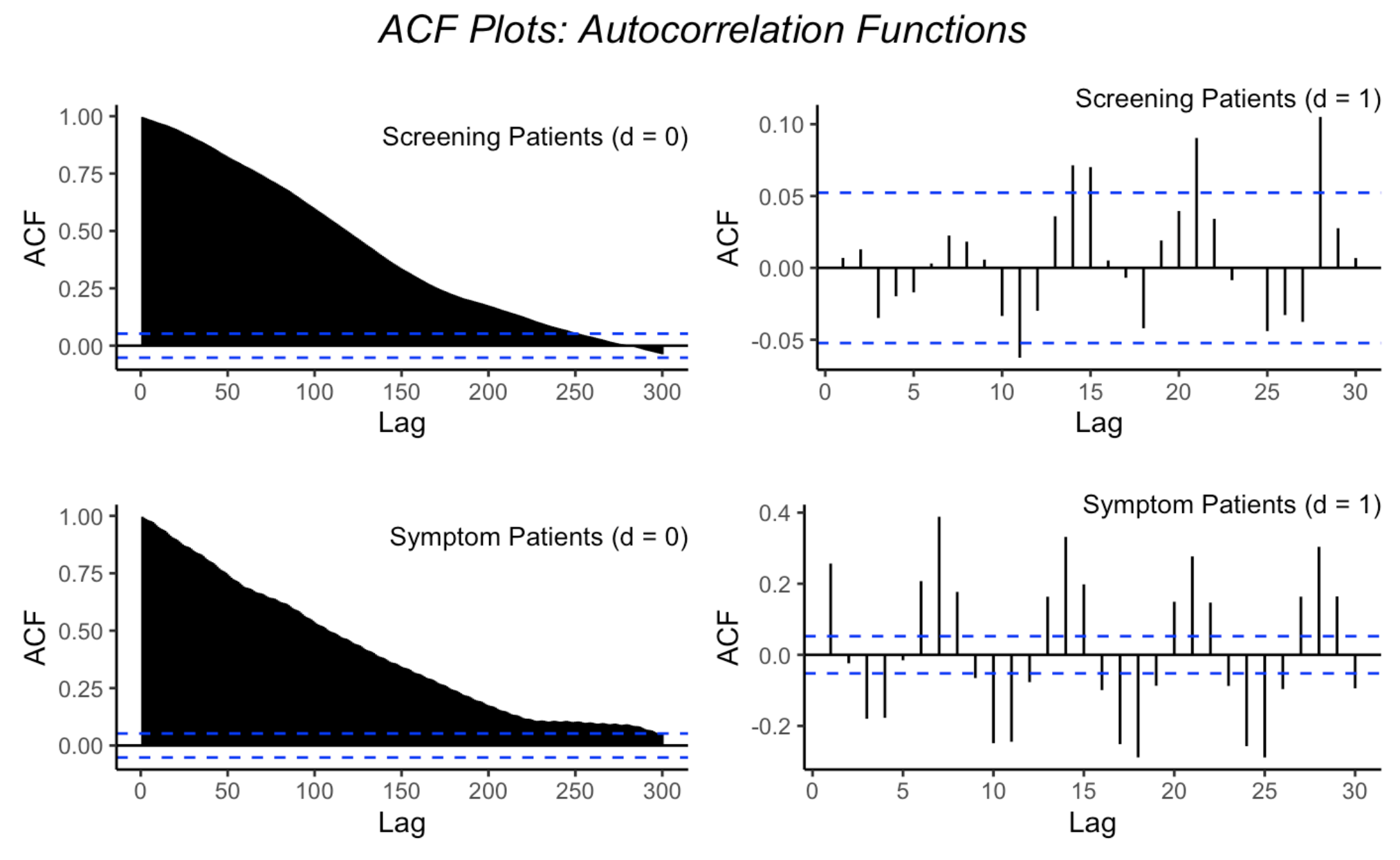

**B. PACF Plots: Partial Autocorrelation Functions**
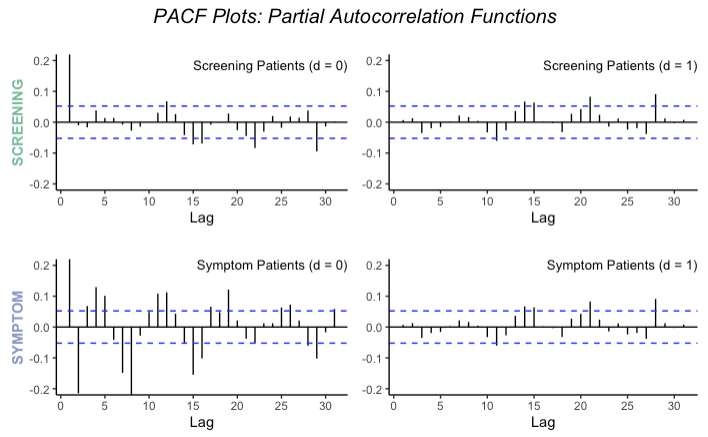

**Figure S12. Distribution of Proportion Delayed**

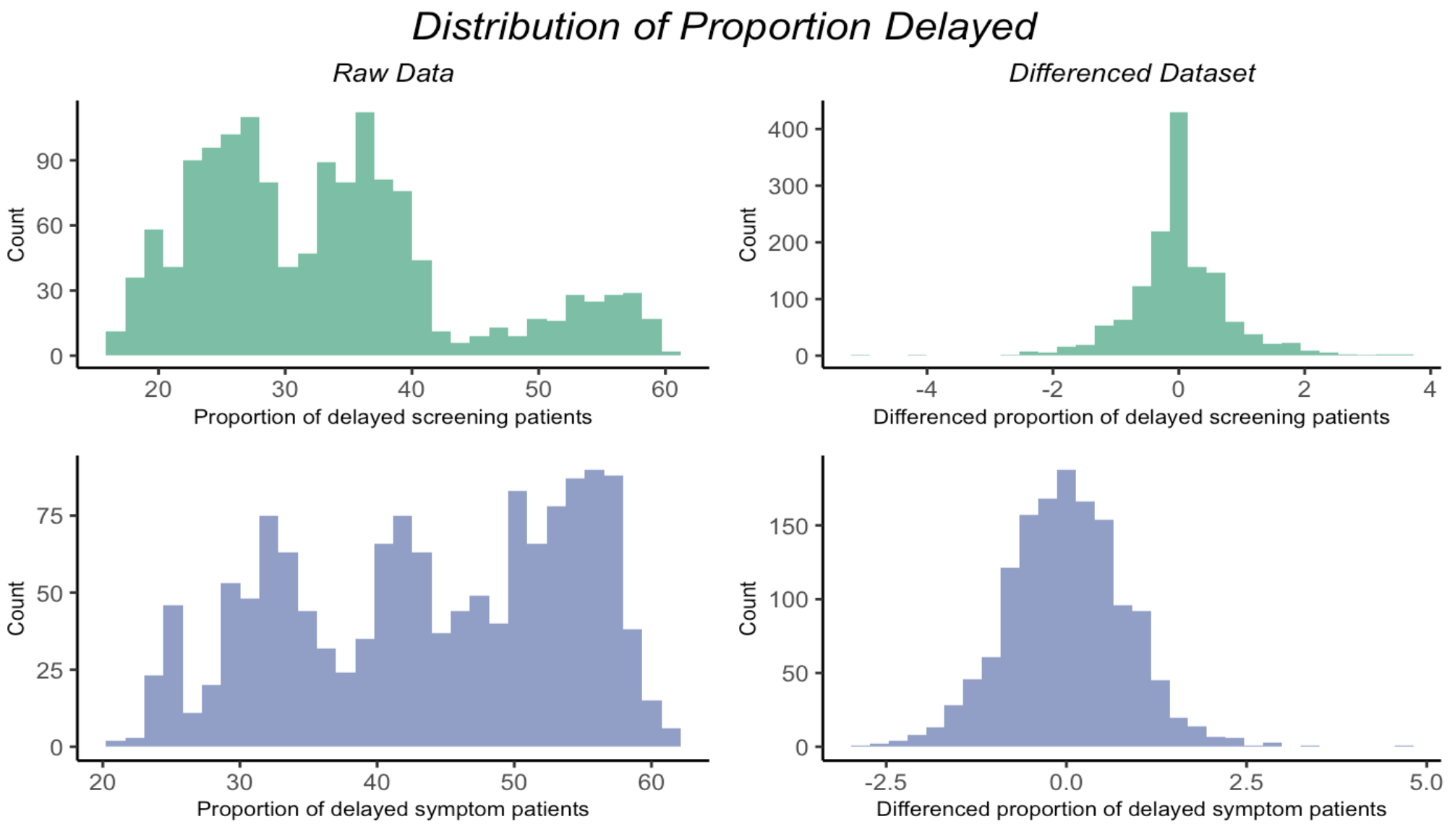

**Figure S13.**  **Characteristic Roots for each SARIMA model.**

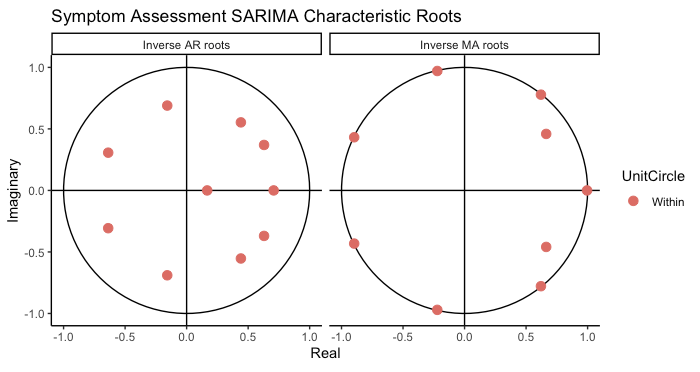

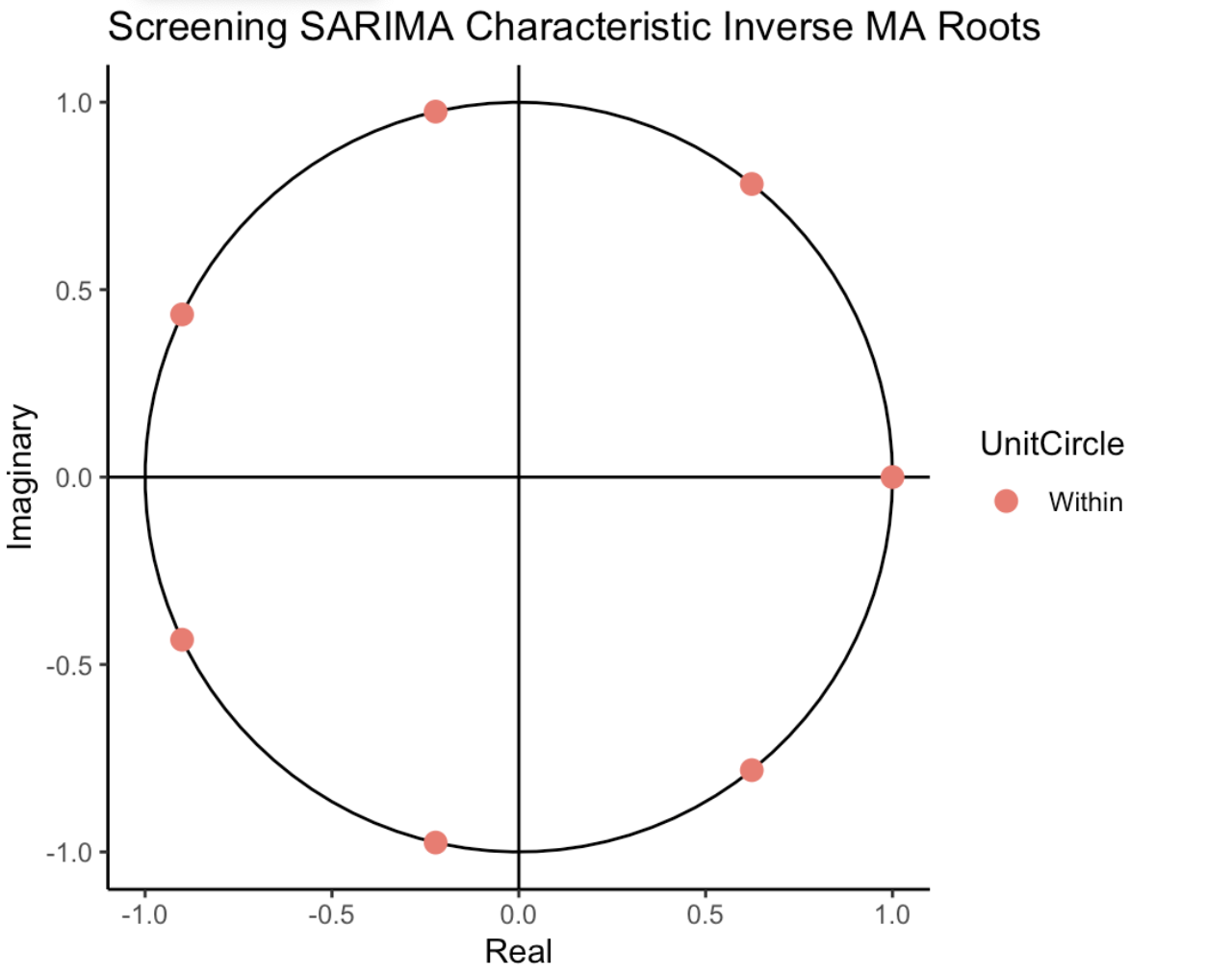

#### Figure S14. Diagnostics Screening SARIMA

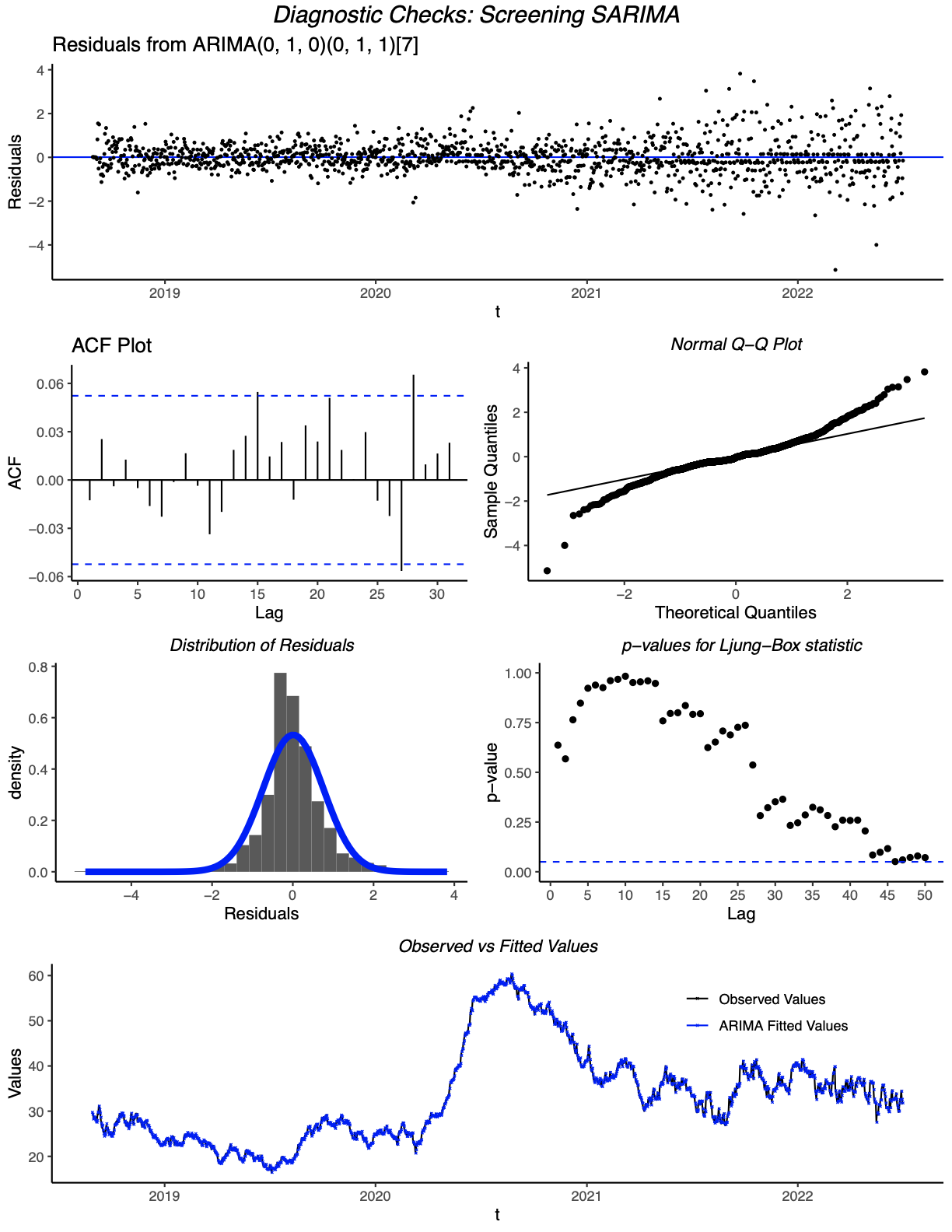

#### Figure S15. Diagnostics Symptom SARIMA

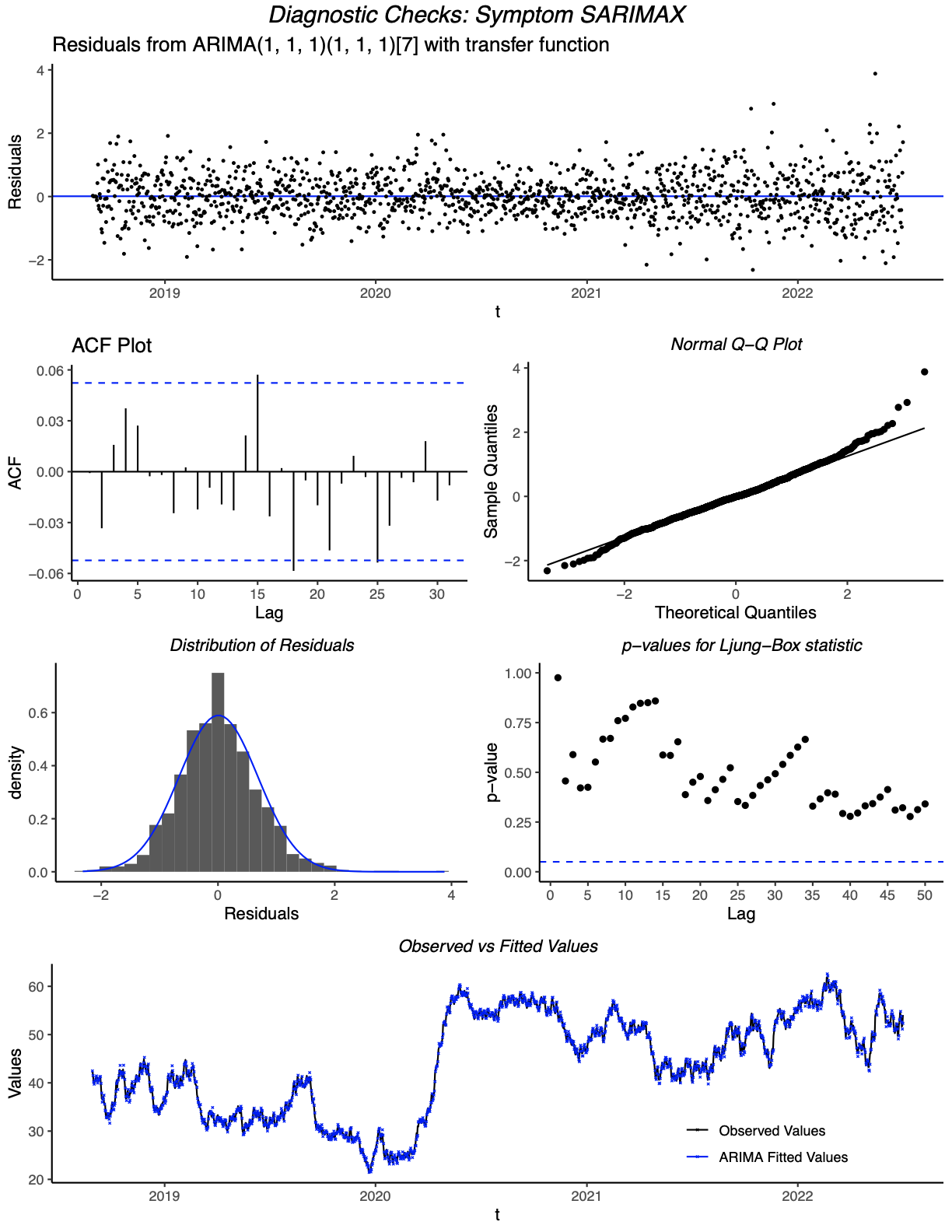

#### Figure S16. Seasonal Window Simulations

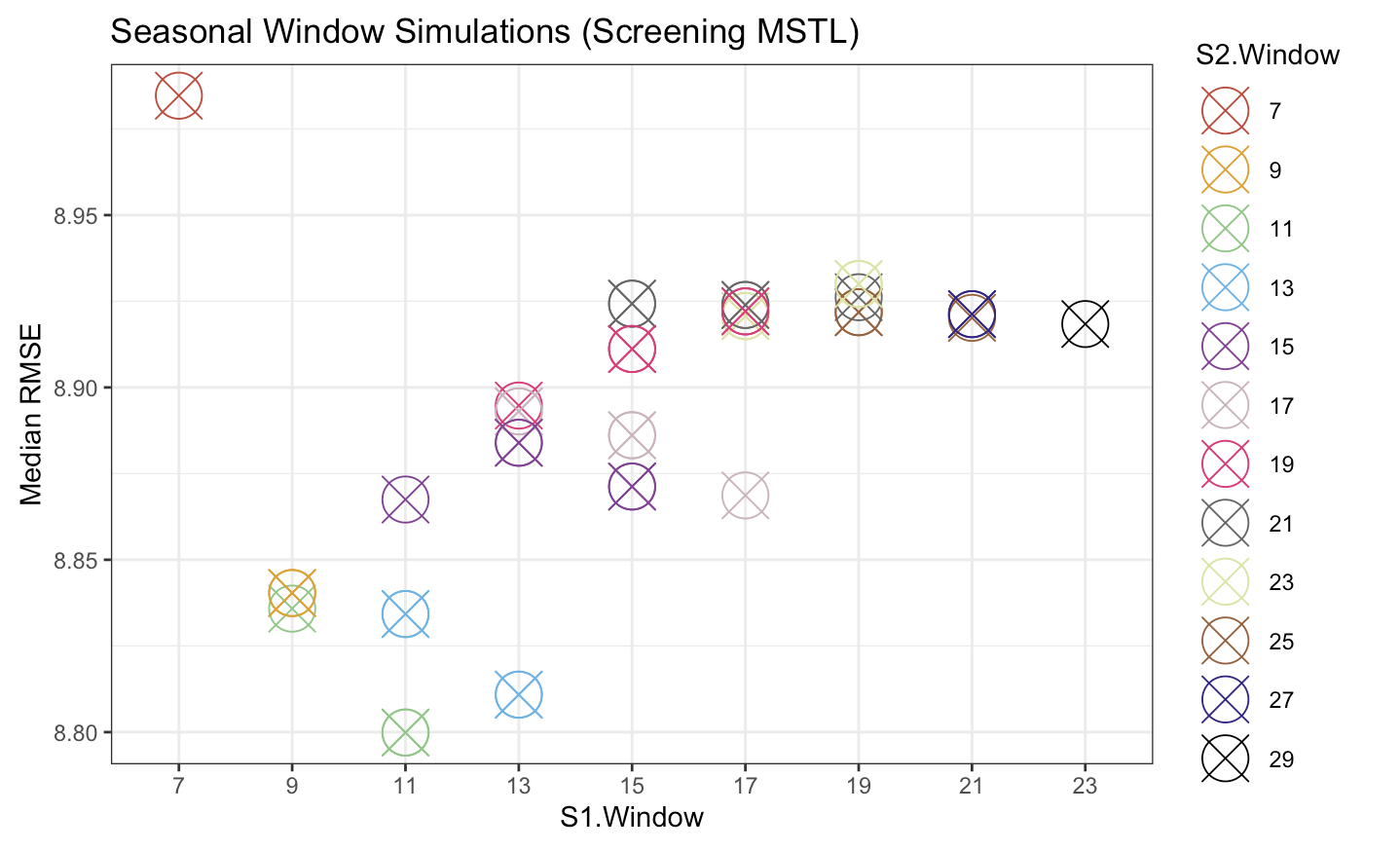

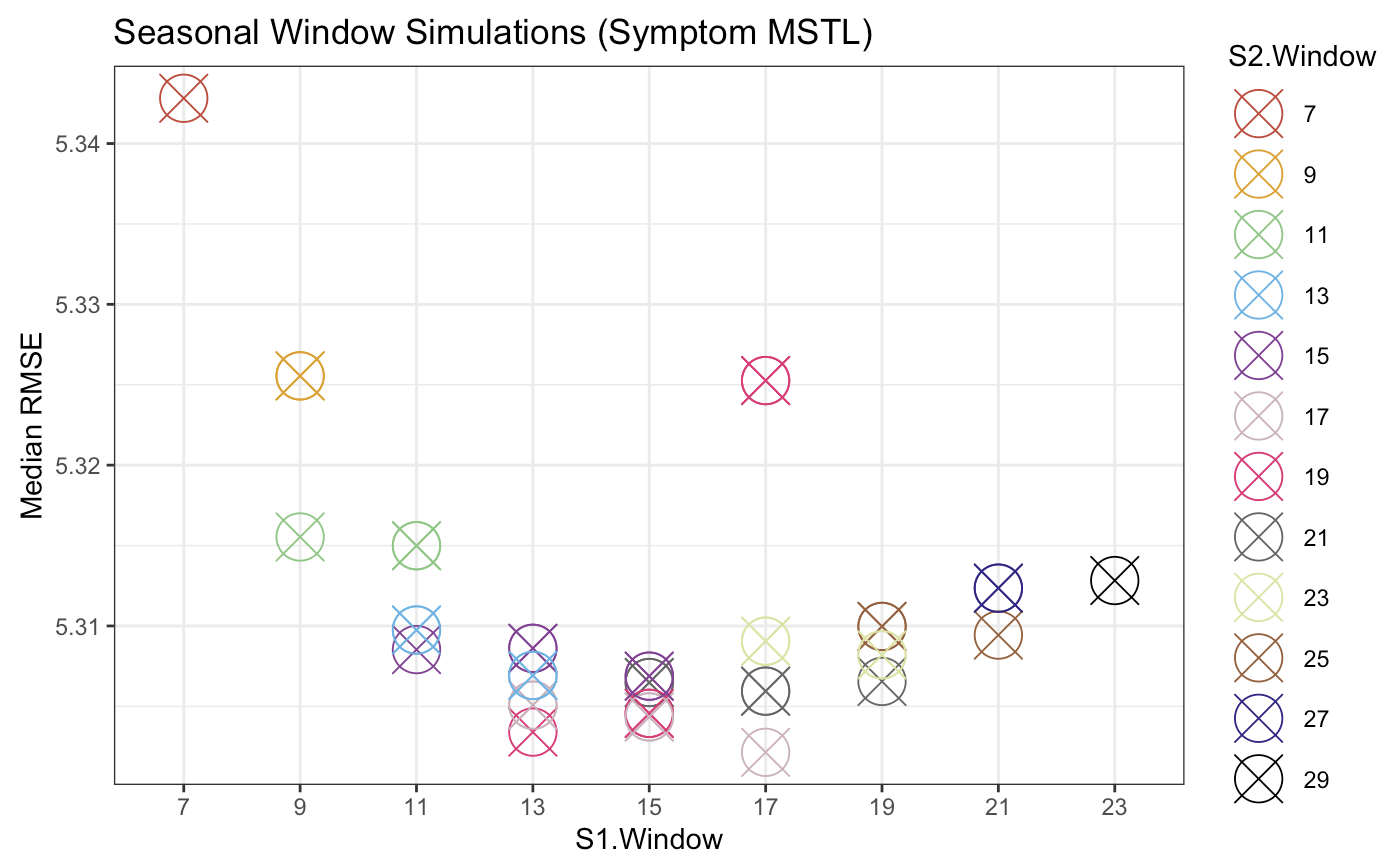
