## Supplement Text for "Impact of COVID-19 Pandemic on Colonoscopy Wait Times by Procedure Indication in Quebec"

#### **Supplement Methods**

#### I. Classification of Patients

We categorized colonoscopy types as “screening”, “surveillance”, or “symptom assessment” based on the indication noted in the CTS **[Figure S1; Table S1]**.

If patient referral forms reported use of anticoagulants, antiplatelets, NSAIDs, insulin, oral anti-diabetic medication and/or having IBD, oxygen-dependent COPD, a cardiac pacemaker, cardiac defibrillation, heart failure, comprehension problems, renal insufficiency, mobility problems, we considered a patient as having a comorbidity. We did not classify patients as having a comorbidity if their referral sheet only indicated “other medication use”.

Screening patients, specifically those without CRC symptoms and at average risk are recommended to undergo an FIT every 2 years after turning 50 years old. Following a positive test, patients are referred for a colonoscopy to confirm the presence of CRC or pre-cancerous adenomas. Of the 2.7 million Quebeckers aged 50-74, 240 727 underwent an FIT in 2022.^1^

Priority 2 patients were considered delayed when their wait time exceeded 2 weeks, priority 3 patients when their time to procedure was over 60 days, and priority 4 patients when their waitlist time was greater than 183 days**[Table S2].** Wait times were longer in the intra-COVID-19 period **[Table S3]**. Among symptom assessment patients (n = 6761), 606 pre-COVID-19 (18.1%) and 1835 intra-COVID-19 (53.8%) patients were deemed to have experienced a delayed procedure **[Table S4].** 178 pre-COVID-19 (12.1%) and 312 intra-COVID-19 (30.7%) screening patients were deemed to have experienced a delayed procedure **[Table S5]**.

A total of 3690 patients were considered to have at least one comorbidity **[Table S6]**. If patient referral forms reported use of anticoagulants, antiplatelets, NSAIDs, insulin, oral anti-diabetic medication and/or having IBD, oxygen-dependent COPD, a cardiac pacemaker, cardiac defibrillation, heart failure, comprehension problems, renal insufficiency, mobility problems, we considered a patient as having a comorbidity. We did not classify patients as having a comorbidity if their referral sheet only indicated “other medication use”.

Descriptive analyses did not detect significant differences in the distribution of age or sex present in patient populations with comorbidities pre vs intra-COVID-19 but suggested a minimal decrease in females <40 with comorbidities among surveillance patients post-COVID-19 **[Table S7].**

#### II. Postal Code Linkage

We used the home address postal code indicated on the colonoscopy referral form to label patients with three neighbourhood characteristics: rural status, MDC, and SDC. In Canada, a “0” in the third digit of a postal code indicates a rural address. To derive each patient’s neighbourhood centiles of deprivation, which are area-based geographic indices, we mapped postal codes to longitudinal and latitudinal coordinates using the *sf R package* based on their spatial proximity^2,3^ and then linked them to their geographic MDC and SCD with the *geojsonio package* using the Canadian Index of Multiple Deprivation dataset.^4^ We linked postal codes with unmatched records to the closest spatial districts’ deprivation indices using nearest neighbours matching. For postal codes that have recently undergone a change (common in high density areas), we used the package *ggmap* to map new postal codes (from 2017 onwards) to the geographic coordinates.

Patients with postal codes on the referral forms outside of Quebec were from across Canada (4 from Nova Scotia, 1 from New Brunswick, 5 from British Columbia, and 23 from Ontario). Seven postal codes (n = 24 patients) required nearest neighbour matching to acquire material deprivation indices **[Figure S2]**. ANOVA testing suggested the MDC and SDC differed significantly across patient populations overall **[Table S7]**. Neighbourhood MDC distribution among patients was right skewed, with an MDC median of 35 (IQR 10-66). Neighbourhood SDC showed a bimodal distribution with a median of 56 (IQR 22-79).

#### III. Regression

#### A. Predictors of Delayed Procedure

A cross over interaction is noted between age and COVID-19, whereby the mean age decreases for patients experiencing delays in the intra-COVID-19 period. The opposite is noted for patients without delay, for which the mean age increases. This interaction noted was supported by results from the *epi.interaction()* function, which reported measures of relative excess risk due to interaction (RERI), the proportion of the outcome among those with both exposures that is attributable to their interaction (AP[AB]), and synergy index indicating a negative interaction.^5^

While the age distribution remained comparable across equivalent strata, COVID-19 did influence the average age of colonoscopy patients experiencing delays or not; Screening patients with a positive FIT tended to be younger in both pre- and intra-COVID-19 periods. For patients without a positive FIT, younger patients were more likely to be delayed in the intra-COVID period compared to the pre-COVID period. We used the *EpiR package* for model development and *ggeffects* package to generate interaction plots.^6^

After exploring the relationship between age, FIT, and COVID-19 graphically and with summary measures **[Figure S3].** A 3-way interaction effect was suspected among screening patients. Whereby, patients with positive FIT were on average older and experienced less delayed procedures than patients with negative or no FIT. Overall, older patients tended to experience greater delayed procedures, however, the mean age of patients with delayed procedures was lower than non-delayed procedures among intra-COVID-19 patients with negative or no FIT. AIC criterion decreased with the addition of the 3-way interaction term, indicating a better fit. However, CI intervals were large, and we concluded our study was not powered enough to detect a significant 3-way interaction.

#### B. Predictors of Normalized Wait Times

Given the distributions to be investigated do not take zeros as values, same day colonoscopies (e.g. proportion delayed is zero) were excluded from analysis (n = 21). While discarding zeros may lead to selection bias,^7^ subanalysis revealed these patients may not be true zeroes and instead are an artefact of data collection or entry error. No systematic bias in patient characteristics is noted among these 21 patients.

Same day colonoscopies (e.g. proportion delayed is zero) were excluded from analysis (n = 21) **[Table S8]**. While discarding zeros may lead to selection bias,^7^ sub analyses revealed these patients may not be true zeroes and instead are an artefact of data collection or entry error. No systematic bias in patient characteristics is noted among these 21 patients.

Cullen and Frey graphs for all patients and subdivided by patient categories and COVID-19 period (pre- or intra-) indicated possible choice of lognormal or gamma distributions for this continuous, non-negative, positively skewed data **[Figure S4]**.

Analyzing goodness-of-fit plots for lognormal, gamma, and Weibull distributions on the dataset revealed that all distributions appropriately describe the centre of the distribution **[Figure S5]**. But the log normal distribution was preferred for its better description of the right tail of the empirical distribution as noted in the Q-Q plot, especially considering this tail is important in the context of delays in access to colonoscopies. Furthermore, the AIC criterion was lowest for the lognormal distribution (17795.61) compared to the gamma and weibull distributions (18875.72 and 18736.83 respectively). The log normal distribution remains the best fit for the skewness/kurtosis of the distribution even when stratified per colonoscopy category**.**

The selected model for predicting log-transformed normalized wait times was a logarithmic linear regression model with covariates age, colonoscopy category, sex, SDC, MDC, COVID-19 period (pre- or intra-), and an interaction term to adjust for the effect modification of age pre/intra-COVID-19. The log normal distribution was preferred for its better description of the right tail of the empirical distribution**.**^7–9^

Our stepwise selection revealed several covariates of interest, which were investigated using univariate log-normal regression. The resulting nested models were compared using ANOVA, which suggested the inclusion of FIT as a significant predictor of decreased wait time.

For diagnostic checks, the residuals histogram showed a normal distribution, therefore the assumption of the normality of residuals is met **[Figure S6-S8]**. There is minimal skew, but sub-analyses reveal this is the result of next day colonoscopies, which, when excluded, do not impact the interpretation of linear regression coefficients. Furthermore, for large sample sizes such as the one included in this paper, the central limit theorem posits the distribution of sample means will follow an approximate normal distribution.^10^

Mixed modelling with random intercepts did not remove clustering among fitted values **[Figure S8]**. We assumed independence, such that each participant’s outcome would be independent from those of the other participants. The consequences of ignoring clustering include increased type 1 error rates.^11^ Additionally, points plotted in diagonal lines among residuals were noted within residual plots, in particular for patients with small wait times; this occurs when a dependent variable is constant or is exactly linearly dependent on predictor variables. Future analyses will investigate more complex model parameters for clustering and non-linear relationships. Given the descriptive nature of the project and diagnostics, the model was deemed sufficient for interpretation.

Two outliers were identified using a histogram plot (proportion delayed > 25) and removed.

Global trends noted in the log-normal linear regression model were the same among symptom and screening patients when analyzed separately **[Table S9]**, however certain patient-type-specific patterns emerged. Residual plots for both patient populations showed similar clustering as noted in the fully adjusted model but met all other assumptions for linear regression.

#### IV. Time Series Analysis

If a random variable *X* is indexed to time, denoted by *t,* the observations *{Xt , t ∈ T}* is called a time series, where T is a time index set (often an integer set). This time index is the time series data type’s main characteristic, usually an equally spaced time interval.^12^ Its analysis serves as a method for understanding the past in order to predict future outcomes. For this report, daily observations were used as discrete time points for analysis.

*X = Proportion delayed of wait list*

*t = Days since start date*

We conducted time series analyses for the proportion of patients on the waitlist classified as delayed (i.e., patients whose wait time exceeded the recommended time interval noted on their referral form).

Times series possesses several features:^13^

- **Cycle/period**: A repeating time interval divides the series into equally long subsets. We used a daily series for a full cycle of 1 year.
- **Frequency**: The frequency is the length or number of units within a cycle. Given this daily series, the frequency was set to 365.25.
- **Timestamp:** The time at which the series was captured. Typically, this time difference (delta) between observations is identical (e.g. daily counts). Thus, the delta was equal to 1 day.

An important characteristic of time series, in particular for ARIMA modelling, is stationarity, Stationary time series data have statistical properties that do not vary with time,^14^ that is, the time series has a constant mean, finite variance, and an auto-covariance structure independent of time^12,15^ If the probability distribution function of the data points is invariant with time, it is deemed strictly stationary.^12^ If the latter condition is unfulfilled, a common occurrence in dynamic systems, the time series data is deemed weakly stationary. This report will move forward with weak stationarity assumptions.

A time series containing a trend or seasonality component is deemed to be non-stationary. A trend is a linear or non-linear long-term increase or decrease in the time series,^15^ whereas seasonality is a pattern occurring at fixed frequencies in the data.^15^ Therefore, a time series that possesses a trend and/or seasonality is considered to be non-stationary given that these components would affect the mean, variance and other statistical properties at any time index.

Certain analytical methods for time series require deseasonalized time series to obtain stationarity.^16^ Given the impact of COVID-19 pandemic measures on access to healthcare, the notion that current volatile colonoscopy waitlist volumes will revert to the long-run mean or average level of the dataset is a assumption open to debate.This report will explore both ARIMA modelling and an STL decomposition model to examine the different components of this time series data. While ARIMA modelling helps mathematically describe data and aims to capture and predict patterns in time-ordered data for forecasting, STL is a decomposition technique used to understand and visualize the underlying structure of the time series.

#### A. ARIMA Model

This analysis used two separate Autoregressive integrated moving average models (ARIMA) to describe the percent delay of the colonoscopy waitlist per colonoscopy category.

The analysis was restricted to priority 2, 3, and 4 patients (ie. only screening and symptom assessment patient populations). Once a patient exceeded the recommended wait time noted on the referral form for their indication, they were considered delayed at time t until their procedure date, where they exited the waitlist **[Figure S9].**

**Equation S1. ARIMA Equation.** An ARIMA(p,d,q) model is noted by:^12^

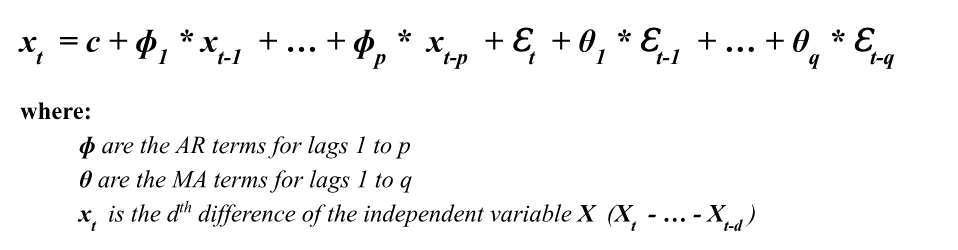

ARIMA models explain time series data based on its past values given by lags and lagged errors.(Nau 2020a) ARIMA models have three components:

1) **Autoregressive (AR)** component models the dependency between the current data point and the previous values in a time series.^14^ The ‘*p*’ parameter represents the order of the AR component, which denotes how many past time steps are used to forecast the current value in the model**.**
2) **Integrated (I)** component represents the number of differencing operations required to make a time series stationary.^14^ This involves subtracting the value of the series at time *t* from a lagged value at time *t-k*. The ‘*d*’ parameter indicates the order of differencing**.**
3) **Moving average (MA)** component refers to the relationship between the current data point and the residual errors from past predictions.^15^ The ‘*q*’ parameter indicates the order of the moving average part.

Here, the outcome (*X_t_*) per the two ARIMA models described are proportion delayed for screening patients and proportion delayed for symptom patients. We used a daily series for a full cycle of 1 year. Given this daily series, the frequency was set to 365.25 and the delta was equal to 1 day. We estimated model parameters using the ADF test, AutoCorrelation Function (ACF), Partial Autocorrelation Function (PACF) and the *auto.arima()* function from the *Forecast package.* The latter is an automated stepwise algorithm combining unit root tests, minimisation of the AICs, and maximum likelihood estimates to output model parameters that best fit the time series data.^15^ Determining model parameters is an iterative process **[Figure S10].**

We assessed the stationarity of the series using ACF plots, together with the ADF and KPSS tests. We used an ACF plot to observe if the process depicted mean reversion behaviour. The autocorrelation functions correlate the delayed variable at time *t* and at time *t-k*. The ADF test is a unit root test where the null hypothesis assumes the presence of a unit root, which is a measure of stochasticity of the model.^17,18^ We conducted a Kwiatkowski-Phillips-Schmidt-Shin (KPSS) test, which is similarly a type of Unit root test but assumes stationarity of a given series around a deterministic trend. In the ACF plot, stationarity was accepted to be true if correlation between time series points across successive lags rapidly approached zero.^19^ A p-value less than 0.05 on the ADF test was deemed significant and indicative of stationarity. Whereas, a p-value less than 0.05 on the KPSS test was deemed significant and indicative of non-stationarity.^20^

An ARIMA model often includes the use of differencing (*d*) of raw observations to induce stationarity into the series data. Differencing helps remove temporal dependence by subtracting the value of a time index from its previous or lagged value^15^ (e.g. differencing takes the differences between successive data points). Slow decay in an ACF plot indicates possible long range dependence or non-stationarity.^21^ We used *diff()* in R to compute the series of changes from one consecutive period to the next, This transforms the data, conferring stationary, as given error terms are assumed to have constant means with finite variance.^22^ We determined the order of differencing based on the significance of the correlations noted in the ACF/PACF and confirmed by the auto.arima() function. We confirmed stationarity by re-running the KPSS and ADF test on the differenced time series.

Further transformations (Box Cox or logarithmic) may be used on differenced data if modelling requirements are unfulfilled (eg. heteroskedasticity and/or a non-normal distribution).^23^ We inspected differenced histograms to confirm a normal distribution and appropriateness for ARIMA modelling. The model is said to take into account the autoregressive structure of the series data, that is, the dependent relationship between an observation and some number of lagged observations.^15^ We determined the number of lagged observations (*p*), also denoted as the lag order, using the ACF and PACF plots.

The ACF and PACF similarly determined the size of the moving average window (*q*). The ARIMA model uses the dependency noted between observations and residual errors from a moving average (MA) model and applies them to a set of lagged observations.^12^ MA models are a linear combination of past random shocks in the model, deemed to have a limited influence on future periods.^14^ Given the different COVID-19 restrictions, at least one of the time series is likely correlated to the errors of previous data points and so will likely be regressed using past forecast errors.

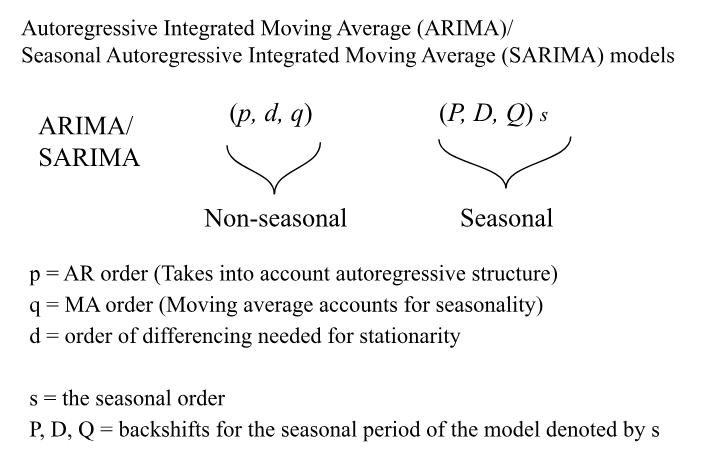

**[**Reproduced from [19]^19,24^ **]**

As predicted by the normal working hours of a hospital (e.g. routine colonoscopies and elective endoscopies tend not to be conducted outside of normal hospital working hours), this potential seasonal element of the time series data needed to be explicitly modelled in a Seasonal ARIMA (SARIMA).^19^ SARIMA models contain the same three parameters as an ARIMA model (p, q, and d), but also have corresponding parameters applied at the seasonal level (P, D, and Q). We determined the seasonal parameters of the model (P, Q, D), using the *auto.arima()* function together with the ACF and the PACF of the differenced time series. Notably, a significant spike at the seasonal lags suggests seasonality. When negative, this spike suggests a seasonal moving average term and when positive suggests a seasonal autoregressive term.^25^ Furthermore, we used a Ljung-box test (h=20)^12^ to assess for autocorrelation in the residuals of each ARIMA/SARIMA model.

The final ARIMA/SARIMA model parameters and graphics help parse out overall trends in the evolution of the proportion of individuals experiencing delays in access to colonoscopies over the COVID-19 pandemic, as well as, how pandemic measures impacted long-term backlogs across screening and symptom assessment patient populations.

For screening patients' waitlists, both the stepwise and non-stepwise non-seasonal model selection algorithms resulted in the same model with parameters (0, 1, 0). The Ljung-Box test was significant (p < 0.05), suggesting the residuals of this model were correlated. For the fitted SARIMA model with weekly seasonality, the non-stepwise algorithm selected SARIMA (0, 1, 0) x (2, 0, 0)[7].

The screening SARIMA model with the lowest BIC information criteria was (0, 1, 0) x (0, 1, 1) [7]. Meaning, the model included first-order differencing (*d = 1*) and a first-order seasonal difference (*D = 1*) to remove trend and induce stationarity in the time series. Considering there is more than one difference taken, no constant (drift) was included as otherwise it would cause quadratic and/or higher order polynomial trends.^19^ The Ljung-Box test was not significant (p > 0.05), suggesting the residuals of this model are independently distributed. The autocorrelation order of the model (*p*) was 0, the moving average order (*q)* was 0, the autocorrelation order of the seasonal component of the model (*P*) was 0, and the moving average order of the seasonal component (*Q*) was 1. This seasonal moving average term captures the influence of the previous season's error terms (previous 7 days). However, the SARIMA model did not include any autoregressive terms.

We performed diagnostic checks on the fitted model to ensure the appropriateness of model parameters, including a plot of the residuals, ACF, and Q-Q plots to check for significant autocorrelation, obvious patterns, and non-normally distributed residuals **[Figure S11]**. We used a Ljung-box test to test for autocorrelation in the residuals of each model.^26^ A p-value of less than 0.05 was considered significant.

Diagnostic tests suggested non-stationarity in both the screening and symptom time series (p-value > 0.05 on ADF; p < 0.05 on KPSS). After first-order differencing, diagnostic tests confirmed stationarity in both screening and symptom time series (p < 0.01 on ADF; p > 0.05 on KPSS) **[Figure S11]** However, we noted a residual seasonal component present in both correlograms and concluded the need to use a multiplicative SARIMA to model the datasets.

Differencing (d) transformed the series to fulfil stationarity assumptions **[Figure S12]**. For the purpose of the screening patient model, we used 1st-order differencing (*d=1*) as it removes non-stationary signals present in the screening and symptom patient datasets, which is suitable for ARIMA modelling**.** Differenced data was normally distributed, thereby indicating no need for further transformations (logarithmic or Box-Cox transformations) to achieve stationarity for ARIMA modelling**.**

Lastly, the characteristic roots were plotted for the final ARIMA models selected to ensure that all inverse roots were within the unit circle, indicating that the model is both stationary and invertible. If roots are close to the unit circle, it is possible the corresponding model is numerically unstable.^15^

The inverse AR roots for the symptom assessment SARIMAX model were within the unit circle, confirming the model as stationary [**Figure S13]**. Meanwhile, the inverse MA roots of the seasonal component for both symptom and screening SARIMA models were close to/on the unit circle, indicating that while the models are unlikely to be good for forecasting, the seasonal part of the models were marginally stable.^27^

Given the *auto.arima()* function returned undifferenced seasonal components, we expect that seasonal differencing induced a non-invertible moving average process. However, as Plosser and Schwert note, over-differencing should not significantly impact the accuracy of estimated model parameters and the validity of inferences.(Plosser and Schwert 1977) Indeed, seasonal differencing aims at removing seasonality and we expect that the previous seasonal period [week] *(T - s)* would impact the current seasonal period [week] (*T*) of patient waitlists. As forecasting was not our primary aim and instead the interpretability of this retrospective analysis was our objective, the model parameters were deemed satisfactory. Which was confirmed by the Ljung-Box test, for which both p-values were not significant (p > 0.05), suggesting the residuals are randomly distributed.

#### B. Intervention Analysis (Interrupted Time Series)

Exceptional external events, known as “intervention events,” can affect a time series of interest. Intervention analysis is used to get a quantitative measure of the impact of an intervention event on a time series being studied.(*Time Series Analysis* 2008). Once model selection is completed, intervention impact can be estimated.

Considering the direct and immediate effect of emergency public health measures put in place to control the spread of COVID-19 on colonoscopy services, we added a transfer function to the ARIMA models to better address the structural break and long-run influence on this time series. Given the declaration of a public health emergency in Quebec on March 13th, 2020 was the catalyst to both emergency policies put into effect and widespread public awareness of the pandemic, this date was considered as a possible change point as well as other dates shortly before or after. We confirmed timing of the event using AIC and BIC criterion of the resulting Seasonal Autoregressive Integrated Moving Average with Exogenous Variables (SARIMAX) model. All dates prior to the change point were deemed “pre-intervention” and all dates until the end of the series were considered to be “post-intervention”.

**[Equation S2: Transfer Function (Step Change)**] ^28^

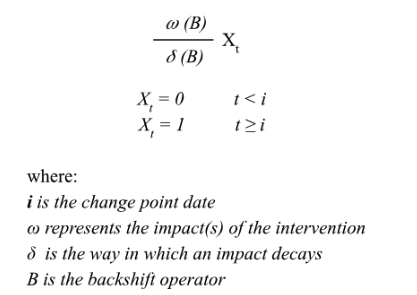

Given trends noted in province-wide data, we assumed there would be an immediate and permanent rise in the proportion of delayed colonoscopies (step change) as well as a change in slope (ramp).(“Intervention Analysis” 1991)

A “ramp” transfer function is a Koyck-type step intervention effect, which is a dynamic response to the intervention at time *t.*(“Intervention Analysis” 1991) For every subsequent period specified (𝛅), there is an added response and each resulting cumulative response is considered permanent.(“Intervention Analysis” 1991)

Variables were included into the model to represent both step and ramp impacts. When calculating the ramp equation, the mean percent difference in the monthly province-wide dataset after March 13th, 2022 was calculated (1.22%) and then approximated per day (0.04%). This was used as the slope of the ramp included per SARIMAX mode land confirmed using AIC/BIC criterion.

A step function was added to both symptom assessment and screening SARIMA models. Several dates were considered, including the 0.01 percentile date of the intra-COVID-19 symptom assessment or screening patients procedure dates in the dataset ("2020-03-16" and “2020-03-19” respectively). The resulting intervention date with the lowest AIC/BIC criterion for symptom assessment patients was March 13th, 2023, when the Quebec Government declared a Public Health Emergency. Meanwhile, for screening patients, the date with the lowest AIC/BIC was when 99.9% of intra-COVID-19 patients procedure dates fell after (“2020-03-19”) **.**

Furthermore, a Ljung-box test (h=20)(Shumway and Stoffer 2017) was used to test if there exists autocorrelation in the residuals of each ARIMAX model.

Finally, intervention analysis allows the estimation of the counterfactual (e.g. a forecast of how the waitlist would have evolved without the interruption of emergency COVID-19 public health measures).(*Time Series Analysis* 2008) The counterfactual was calculated by applying the final ARIMA model to the data pre-intervention.(Schaffer, Dobbins, and Pearson 2021) We then plotted 80 days of predictions post-intervention date against the observed values.

All screening SARIMAX models had an AIC/BIC greater than the initialized screening SARIMA models and so, the SARIMA model was considered the final model for screening. However, log-likelihoods were lower when including a step and ramp variable and the Box-Ljung test were non-significant, suggesting there was no significant autocorrelation among the residuals.

For the screening ARIMA(0, 1, 0)(0, 1, 1)[7] model, the residuals were normally distributed with some minimal left-skew, explained by outliers **[Figure S14]**. While the Ljung-Box test showed non-significance until H = 45, the Q-Q plot of the residuals suggests the model is not capturing all variability within our data. Indeed, greater variation is noted in residuals after 2021. Variability in residuals was not improved with Box Cox (lambda determined by *BoxCox.lambda()*) or logarithmic transformations.

The step SARIMAX model (1, 1, 1)(1, 1, 1)[7] for symptom assessment patients had an AIC and log-likelihood greater than the initial symptom assessment SARIMA models, however it had a lower BIC and less model parameters. Thus, we chose this SARIMAX model as the one that best describes the symptom assessment waitlist. Including a transfer function was further justified by the multiplicative trend noted in the STL model. The Box-Ljung test was non-significant, suggesting there was no significant autocorrelation among the residuals.

For the symptom assessment ARIMAX(1, 1, 1)(1, 1, 1)[7] model, the residuals were normally distributed with some right skew **[Figure S15]**. The skew is accounted for by an outlier. There is a small significant spike at 15 and 18 of the autocorrelation plot, however, the Ljung-Box test showed non-significance with H > 50 and the Q-Q plot of the residuals suggests normal distribution.

C. STL Model

To understand ARIMA modelling results, we further decomposed our time series data using an STL model. STL modelling is a filtering procedure that decomposes time series into trend, seasonal, and remainder components (Petropoulos et al. 2022; Cleveland, Cleveland, and Terpenning 1990). Through this process, researchers can gather insights on the underlying patterns of the data and the behaviour of each component individually.

STL modelling applies a smoothing function on the time series using LOESS, a non-parametric curve-fitting procedure. This process occurs in two iterative processes: an inner loop nested inside an outer loop.(Cleveland, Cleveland, and Terpenning 1990) The inner loop iterates *n_(i)_* times between seasonal and trend smoothing, and the outer loop minimizes the impact of outliers (*n_(o)_* robustness iterations).

STL model parameters provide lots of flexibility and this may help capture a wide range of seasonal fluctuations/patterns.(Cleveland, Cleveland, and Terpenning 1990) The steps of the inner loop is pictured here adapted from [X](Zhang et al. 2023):

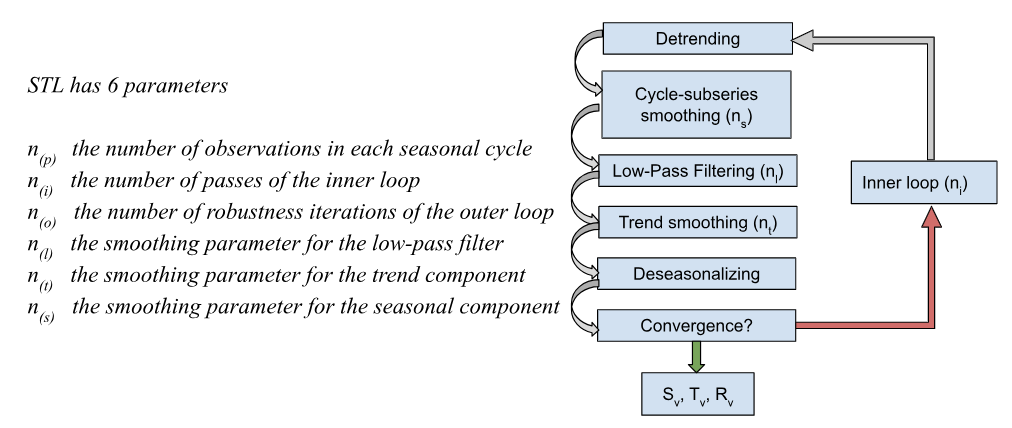

Within the inner loop, the seasonal component is calculated first where LOESS allows the seasonal component to be detrended. The series is then deseasonalized before deriving the trend component. The remainder is then determined by subtracting the seasonal and trend components from the initial time series (deseasonalized and detrended) **[Equation 4]**.

**Equation S3. STL Equation.** The resulting additive decomposition is denoted by: (Bandara, Hyndman, and Bergmeir 2021)

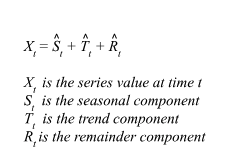

The data was transformed to a multi seasonal time series (msts) object with seasonal periods of 7 days, 30 days, and 366 days to represent weekly, monthly, and annual seasonality. Meaning, n_(p)_ = [7, 30, 366], and n_(l)_ = [n_(p)_]_odd_ (automated by the mstl() function).(Cleveland, Cleveland, and Terpenning 1990; Bandara, Hyndman, and Bergmeir 2021) The presence of three seasonal components was confirmed using seasonal diagnostic plots. We used a robust MSTL model for both screening and symptom assessment populations (n_(i)_ = 2 and n_(o)_ = 10).(Cleveland, Cleveland, and Terpenning 1990)

The resulting screening MSTL showed increased variance around the beginning of the intra-COVID-19 period. Annual seasonality is noted, with peaks in the late summer, early fall. The model has time-varying weekly and monthly seasonal patterns, capturing greater changes and stochastic processes in the proportion delayed of screening colonoscopy waitlists. The trend component indicated a peak in the proportion delayed following the emergency declaration in Quebec, with a gradual return to pre-COVID-19 values, however the proportion delayed still remains greater in 2022 compared to the pre-COVID-19 period.

The symptom MSTL similarly described increased variance around the start of the intra-COVID-19 period. Annual seasonality is not significant, approaching a white noise pattern. The model also has time-varying weekly and monthly seasonal patterns. The trend component indicated a slow decline in proportion delayed pre-COVID-19, however after the emergency declaration in Quebec there is an exponential increase, with a slight plateau before returning to an increased proportion delayed in early 2021.

The yearly seasonality identified in screening patients waitlists, specifically the increased delays in autumn are in accordance with decreases in colonoscopies conducted in summer, as well as, increases in the number of FIT conducted in the spring.(“Cancérologie | Cancer colorectal” n.d.) Work schedules are likely the source of most seasonal components, with regular hospital hours keeping with a 5-day work week schedule and peaks and troughs coinciding with winter and summer holidays.

Lastly, the smoothing parameter for the trend component [n_(t)_] is odd and denoted by 1.5*n_(p)_* / (1 - 1.5/*n_(s)_ ).*(Bandara, Hyndman, and Bergmeir 2021; “6 - Time Series Decomposition” n.d.) We selected the trend component by running the model with the different *n_(p)_* and respective *n_(s)_* per seasonal component and selecting the pattern that minimized residuals but was not subject to over-fitting.

#### Seasonal Window Simulations

We assumed weekly and monthly seasonal patterns evolved quickly (and so were time-varying) due to the changing policy landscape from 2018 to 2022. We determined the rate of seasonal variation or s.window parameters [n_(s)_] for the S1.window (weekly) and S2.window (monthly) using a simulation study as described by Bandara et al.(Bandara, Hyndman, and Bergmeir 2021) We evaluated various combinations of s.window values satisfying the S1.Window < S2.Window condition **[Equation 6]**.

**Equation S4. Seasonal Variation Parameters.** The pairs were generated based on:(Bandara, Hyndman, and Bergmeir 2021)

*where:*

*C = (7, 9, 11, 13, 15)*

*K = (0, 1, 2 ,3 ,4 ,5 ,6 ,7)*

*i = 1 for the first seasonality (weekly)*

*i = 2 for the second seasonality (monthly)*

*S1.Window < S2.Window*

*so:*

*S = (C + K*i, C+K*i +1)*

*(C + K, C + K + 1)_odd_	i=1*

*(C + 2K, C + 2K + 1)_odd_	i=2*

*S1, S2*

We conducted a simulation for screening and symptom assessment patient waitlists separately, selecting the paired S1.Window and S2.Window parameters that yielded the best median RMSE as the default s.window parameter for the MSTL models. We assumed that the annual seasonal pattern was constant over time and so, a deterministic process.

The paired S1.Window and S2.Window yielding the best median RMSE from the simulation study where S1.Window < S2.Window, was selected as the default s.window parameter for the MSTL models **[Figure S16]**. For the screening MSTL, the S1.Window was set to 11 and the S2.Window was 13. For the symptom assessment MSTL, the S1.Window and the S2.Window were n_(S1)_ = 13 and n_(S2)_ = 19 respectively. The S3.Window was set to periodic (n_(S3)_ = 9999). Other parameters explored with a similar simulation study for the annual seasonal pattern (S1.Window < S2.Window < S3.Window) showed no gains in model fit. The trend component selected that minimized the remainder without overfitting the decomposition was based on the monthly seasonal component for both models, with n_(t)_ = 51 for the screening MSTL and n_(t)_ = 49 for the symptom MSTL.
