## Supplement Tables for "Impact of COVID-19 Pandemic on Colonoscopy Wait Times by Procedure Indication in Quebec"

**Table S1. Description of Colonoscopy Categories** **Based on Referral Form**

| **Colonoscopy Category** | *Symptom* | *Screening* | *Surveillance* |
| --- | --- | --- | --- |
| **Indication** | IN2  IN3, IN4, IN6, IN7, IN17, IN18, IN19, IN20  IN10, IN12 | IN5, IN8, IN11 | IN14, IN13, IN15, IN21 |
| **Priority** | 2, 3, 4 | 3, 4, 5 | “Follow-up” |
| **Target Timeline** | Yes | Yes | No |

**[Table S1. Description of Colonoscopy Categories** **Based on Referral Form.** Priority 1 patients (IN1: Acute lower gastrointestinal haemorrhage) were excluded from analysis. IN1: Acute lower gastrointestinal haemorrhage; IN2: High index of suspicion for cancer; IN3: Suspicion for IBD, IN4: Haematochezia ≥ 40 years old; IN5: FIT test; IN6: Iron deficiency anaemia; IN7: Change in bowel habits; IN8: Family history of CRC or polyps; IN10: Haematochezia < 40 years old; IN11 Screening for CRC with average risk; IN12: Chronic constipation or diarrhoea; IN13: History of polyps; IN43: History of CRC; IN15: History of IBD; IN17: Polyps on imagine; IN18: Suspicion of CRC from paraneoplastic syndrome; IN19: Inadequate bowel prep; IN20: Post-acute diverticulitis; IN21: Significant family history]

**Table S2. Delays Based on Patient Priority.**

| **Priority** | **Target Timeline** | **Indication** |
| --- | --- | --- |
| 2 | Less than 14 days | IN2 |
| 3 | Less than 60 days | IN3, IN4, IN5, IN6, IN7, IN17, IN18, IN19, IN20 |
| 4 | Less than 6 months (183 days) | IN8, IN10, IN12 |
| 5 (noncompliant screening) | N/A | IN11 |
| “Follow-up” (surveillance) | N/A | IN14, IN13, IN15, IN21 |

**[Table S2. Delays Based on Patient Priority.** Indications have different priorities with a recommended maximum wait time for colonoscopy procedure. Priority 5 and Follow-up patients have no recommended timeline and so cannot be characterized with a binary indicator of delayed or not. We excluded these patients whose delay cannot be determined from the CTS form from our analysis of delay. Indeed, patients referred to colonoscopy for colorectal cancer screening without family history or a positive FIT (Priority 5) have no target timeline due to non-compliance with screening guidelines. Meanwhile, surveillance patients have wait times based on time since their last procedure which was not in our dataset.]

#### **Table S3. Sociodemographic Variables and Colonoscopy Findings for Surveillance Patients**

| **Surveillance Patients** | *Pre-COVID*  (n = 2231) | *Intra-COVID*  (n = 2425) | *p-value* |
| --- | --- | --- | --- |
| ***Age (years)*** |  |  | **0.002^2^** |
| Mean (SD) | 62.39 (12.24) | 63.49 (12.10) |  |
| Range | 18.00-95.00 | 18.00-93.00 |  |
| ***Sex*** | |  | 0.835^1^ |
| Female | 1042 (46.7%) | 1140 (47.0%) |  |
| Male | 1189 (53.3%) | 1285 (53.0%) |  |
| ***BMI*** | |  | 0.164^2^ |
| N-Miss | 551 | 623 |  |
| Mean (SD) | 26.91 (5.13) | 27.15 (5.15) |  |
| ***Rural*** | |  | 0.930^1^ |
| Rural | 95 (4.3%) | 102 (4.2%) |  |
| Urban | 2136 (95.7%) | 2323 (95.8%) |  |
| ***Material Deprivation Centile*** | |  | 0.504^2^ |
| Mean (SD) | 38.59 (30.73) | 39.20 (31.27) |  |
| ***Social Deprivation Centile*** | |  | 0.078^2^ |
| Mean (SD) | 48.88 (31.60) | 47.26 (31.24) |  |
| ***Time to Procedure*** | |  | **< 0.001^2^** |
| Mean (SD) | 130.40 (159.00) | 308.24 (347.40) |  |
| ***Cancer*** | |  | 0.325^1^ |
| Prevalence | 8 (0.4%) | 5 (0.2%) |  |
| ***Clinical Significant Lesions*** | |  | 0.886^1^ |
| Prevalence | 1059 (47.5%) | 1146 (47.3%) |  |

*SD, Standard Deviation; 1. Pearson's Chi-squared test; 2. Linear Model ANOVA*

#### **Table S4. Sociodemographic Variables and Colonoscopy Findings for Symptom Patients**

| **Symptom Patients** | *Pre-COVID*  (n = 3351) | *Intra-COVID*  (n = 3410) | *p-value* |
| --- | --- | --- | --- |
| ***Age (years)*** |  |  | **<0.001^2^** |
| Mean (SD) | 55.71 (15.48) | 57.15 (15.42) |  |
| Range | 18.00-95.00 | 18.00-99.00 |  |
| ***Sex*** | |  | 0.356^1^ |
| Female | 1811 (54.0%) | 1881 (55.2%) |  |
| Male | 1540 (46.0%) | 1529 (44.8%) |  |
| ***BMI*** | |  | 0.664^2^ |
| N-Miss | 792 | 895 |  |
| Mean (SD) | 26.62 (5.69) | 26.69 (5.53) |  |
| ***Rural*** | |  | **0.029^1^** |
| Rural | 119 (3.6%) | 157 (4.6%) |  |
| Urban | 3232 (96.4%) | 3253 (95.4%) |  |
| ***Material Deprivation Centile*** | |  | 0.643^2^ |
| Mean (SD) | 40.39 (31.84) | 40.03 (31.52) |  |
| ***Social Deprivation Centile*** | |  | 0.132^2^ |
| Mean (SD) | 53.42 (31.83) | 52.26 (31.53) |  |
| ***Time to Procedure*** | |  | **< 0.001^2^** |
| Mean (SD) | 46.48 (59.39) | 97.05 (118.12) |  |
| ***Procedure delayed based on triage reference sheet*** | | | **< 0.001^1^** |
| Delayed | 606 (18.1%) | 1835 (53.8%) |  |
| On-time | 2745 (81.9%) | 1573 (46.2%) |  |
| ***Cancer*** | |  | 0.941^1^ |
| Prevalence | 35 (1.0%) | 35 (1.0%) |  |
| ***Clinical Significant Lesions*** | |  | **< 0.001^1^** |
| Prevalence | 962 (28.7%) | 1180 (34.6%) |  |

*SD, Standard Deviation; 1. Pearson's Chi-squared test; 2. Linear Model ANOVA*

#### **Table S5. Sociodemographic Variables and Colonoscopy Findings for Screening Patients**

| **Screening Patients** | *Pre-COVID*  (n = 1856) | *Intra-COVID*  (n = 1287) | *p-value* |
| --- | --- | --- | --- |
| ***Age (years)*** |  |  | **0.013^2^** |
| Mean (SD) | 59.34 (11.39) | 60.36 (11.22) |  |
| Range | 18.00-93.00 | 18.00-91.00 |  |
| ***Sex*** | |  | 0.230^1^ |
| Female | 897 (48.3%) | 650 (50.5%) |  |
| Male | 959 (51.7%) | 637 (49.5%) |  |
| ***BMI*** | |  | **0.010^2^** |
| N-Miss | 408 | 301 |  |
| Mean (SD) | 26.50 (5.12) | 27.06 (5.36) |  |
| ***Rural*** | |  | 0.508^1^ |
| Rural | 57 (3.1%) | 45 (3.5%) |  |
| Urban | 1799 (96.9%) | 1242 (96.5%) |  |
| ***Material Deprivation Centile*** | |  | 0.714^2^ |
| Mean (SD) | 38.41 (31.54) | 38.58 (31.30) |  |
| ***Social Deprivation Centile*** | |  | **0.051**^2^ |
| Mean (SD) | 52.59 (31.82) | 50.34 (31.44) |  |
| ***Time to Procedure*** | |  | **< 0.001^2^** |
| Mean (SD) | 74.84 (83.40) | 140.03 (197.89) |  |
| ***Procedure delayed based on triage reference sheet*** | | | **< 0.001^1^** |
| N-Miss (Priority 5) | 391 | 274 |  |
| Delayed | 178 (12.2%) | 312 (30.8%) |  |
| On-time | 1287 (87.8%) | 701 (69.2%) |  |
| ***Cancer*** | |  | 0.664^1^ |
| Prevalence | 24 (1.3%) | 19 (1.5%) |  |
| ***Clinical Significant Lesions*** | |  | 0.682^1^ |
| Prevalence | 804 (43.3%) | 567 (44.1%) |  |

*SD, Standard Deviation; 1. Pearson's Chi-squared test; 2. Linear Model ANOVA*

#### **Table S6. Patients with Comorbidities**

| *Patients with Comorbidities* | **n = 3683** |
| --- | --- |
| ***Medication Use*** | **n = 2649 (72%)** |
| Anticoagulants | 583 (15.8%) |
| Antiplatelets | 1090 (29.6%) |
| NSAIDs | 285 (7.7%) |
| Insulin | 316 (8.6%) |
| Oral Anti-Diabetic Rx | 1139 (30.9%) |
| Other | 2969 (n/a) |
| ***Pathology*** | **n = 1379 (37%)** |
| IBD | 924 (25.0%) |
| Oxygen-Dependent COPD | 17 (0.5%) |
| Cardiac Pacemaker | 53 (1.4%) |
| Cardiac Defibrillation | 19 (0.5%) |
| Heart Failure | 9 (0.2%) |
| Comprehension Problems | 70 (1.9%) |
| Mobility Problems | 100 (2.7%) |
| Renal Insufficiency | 285 (7.7%) |

*NSAID, nonsteroidal anti-inflammatory drug; Rx, Prescription; IBD, Inflammatory Bowel Disease; COPD, chronic obstructive pulmonary disease*

#### **Table S7. Socodemographic Variables and Colonoscopy Findings for Urban vs Rural Patients**

| ***Socio-demographic Variables for Urban vs Rural Patients*** | | | |
| --- | --- | --- | --- |
| **Geographic Location** | *Rural*  (n = 575) | *Urban*  (n = 13985) | *p-value* |
| ***Age (years)*** |  |  | 0.791^2^ |
| Mean (SD) | 59.39 (13.10) | 59.23 (13.97) |  |
| Range | 18.00-88.00 | 18.00-99.00 |  |
| ***Sex*** | |  | 0.492^1^ |
| Female | 285 (49.6%) | 7136 (51.0%) |  |
| Male | 290 (50.4%) | 6849 (49.0%) |  |
| ***BMI*** | |  | 0.069^2^ |
| N-Miss | 166 | 3404 |  |
| Mean (SD) | 27.27 (5.46) | 26.77 (5.38) |  |
| ***COVID-19*** | |  | 0.053^1^ |
| Pre | 271 (47.1%) | 7167 (51.2%) |  |
| Intra | 304 (52.9%) | 6818 (48.8%) |  |
| ***Material Deprivation Centile*** | |  | **<0.001**^2^ |
| Mean (SD) | 61.00 (32.46) | 38.55 (31.03) |  |
| ***Social Deprivation Centile*** | |  | **<0.001**^2^ |
| Mean (SD) | 38.21 (25.26) | 51.57 (31.80) |  |
| ***Time to Procedure*** | |  | 0.710^2^ |
| Mean (SD) | 129.70 (206.46) | 126.54 (199.55) |  |
| ***Cancer*** | |  | 0.165**^1^** |
| Prevalence | 8 (1.4%) | 118 (0.8%) |  |
| ***Clinically Significant Lesions*** | |  | **0.003**^1^ |
| Prevalence | 260 (45.2%) | 5458 (39.0%) |  |

*SD, Standard Deviation; 1. Pearson's Chi-squared test; 2. Linear Model ANOVA*

**Table S8. Same-Day Colonoscopies**

| *Same Day Colonoscopies* | **n = 21** |
| --- | --- |
| ***COVID-19*** |  |
| Pre | 9 |
| Intra | 12 |
| ***Colonoscopy Category*** |  |
| Screening | 3 |
| Symptom | 18 |
| ***Sex*** |  |
| Female | 9 |
| Male | 12 |
| ***Deprivation Indices (median)*** |  |
| Material | 28 |
| Social | 74 |
| ***Age*** |  |
| Median | 59 |
| Range | 18 - 85 |

| Table S9. **Base Logistic Regression Model for Delayed Procedures for Screening and Symptom Patients** | | | | |
| --- | --- | --- | --- | --- |
|  | *Screening (n = 2475)* | | *Symptom (n = 6741)* | |
| **Covariate** | **Adjusted OR ^1^**  **(95% CI)** | **p-value** | **Adjusted OR ^1^ (95% CI)** | **p-value** |
| *COVID-19* |  |  |  |  |
| Pre | Ref. | - | Ref. | - |
| Intra | 3.58 (2.90, 4.44) | **<0.001** | 5.26 (4.71-5.88) | **<0.001** |
| *Sex* |  |  |  |  |
| Male | Ref. | - | Ref. | - |
| Female | 1.00 (0.81, 1.23) | 0.988 | 1.11 (1.00-1.24) | **0.048** |
| *Age ^2^* |  |  |  |  |
| Per decade | 1.03 (0.94, 1.13) | 0.489 | 1.05 (1.02-1.09) | **0.003** |
| *FIT* |  |  |  |  |
| Negative or  No FIT | Ref. | - |  |  |
| Positive FIT | 0.56 (0.45, 0.70) | **< 0.001** |  |  |
| *Deprivation Dentile ^2^* |  |  |  |  |
| Material | 1.00 (0.97, 1.03) | 0.951 | 0.99 (0.97, 1.01) | 0.232 |
| Social | 1.02 (0.99, 1.06) | 0.149 | 1.01 (0.99, 1.02) | 0.487 |

*OR, Odds ratio; CI, Confidence Interval; Ref. referent category*

*^1^ Exponentiated regression coefficient*

*^2^ Covariates were rescaled by 10*

**Table S10. Diagnostic results for candidate ARIMA/SARIMA models for Proportion of Waitlist Delayed for Screening Patients (selected model is bolded)**

| ***ARIMA and SARIMA models for Screening Patients*** | | | | | | | |
| --- | --- | --- | --- | --- | --- | --- | --- |
| **ARIMA/**  **SARIMA** | **Non-seasonal Terms**  **(p, d, q)** | **Seasonal**  **Terms**  **(P, D, Q)** | **Seasonal Order**  **(s)** | **AIC/BIC** | **σ^2^** | **Log-**  **likelihood** | **p-value of Ljung Box test** |
| ARIMA stepwise  auto.arima() | (0, 1, 0) | (0, 0, 0) | 0 | 3238.95 / 3244.2 | 0.59 | -1618.48 | 0.026 |
| ARIMA non-stepwise  auto.arima() | (0, 1, 0) | (0, 0, 0) | 0 | 3238.95 / 3244.2 | 0.59 | -1618.48 | 0.026 |
| SARIMA non-stepwise  auto.arima() | (0, 1, 0) | (2, 0, 0) | 7 | 3234.9 / 3250.71 | 0.59 | -1614.48 | 0.30 |
| **SARIMA arima()** | **(0, 1, 0)** | **(0, 1, 1)** | **7** | **3199.60 / 3210.08** | **0.56** | **-1597.8** | **0.79** |
| SARIMA arima() | (0, 1, 0) | (1, 0, 1) | 7 | 3199.21 / 3214.94 | 0.57 | -1596.6 | 0.74 |
| SARIMA arima()  [with step] | (0, 1, 0) | (0, 1, 1) | 7 | 3200.90 / 3216.62 | 0.56 | -1597.5 | 0.78 |
| SARIMA arima()  [with step and ramp] | (0, 1, 0) | (0, 1, 1) | 7 | 3202.10 / 3223.06 | 0.56 | -1597.05 | 0.78 |

**Table S11. Diagnostic results for candidate ARIMA/SARIMA models for Proportion of Waitlist Delayed for Symptom Patients**

| ***ARIMA and SARIMA Models for Symptom Patients*** | | | | | | | |
| --- | --- | --- | --- | --- | --- | --- | --- |
| **ARIMA/**  **SARIMA** | **Non-seasonal Terms**  **(p, d, q)** | **Seasonal Terms**  **(P, D, Q)** | **Seasonal Order**  **(s)** | **AIC/BIC** | **σ^2^** | **Log-**  **likelihood** | **p-value of Ljung Box test** |
| ARIMA stepwise  auto.arima() | (0, 1, 5) | (0, 0, 0) | 0 | 3330.23 / 3361.71 | 0.63 | -1659.12 | < 0.001 |
| ARIMA non-stepwise  auto.arima() | (5, 1, 0) | (0, 0, 0) | 0 | 3303.57 / 3335.05 | 0.61 | -1645.78 | < 0.001 |
| SARIMA non-stepwise auto.arima() | (1, 1, 0) | (2, 0, 0) | 7 | 3127.79 / 3148.77 | 0.54 | -1559.89 | < 0.001 |
| SARIMA arima() | (5 , 1, 0) | (0, 1, 1) | 7 | 2912.99 / 2949.68 | 0.46 | -1449.5 | 0.33 |
| SARIMA arima() | (4, 1, 1) | (0, 1, 1) | 7 | 2911.51 / 2948.2 | 0.46 | -1448.76 | 0.42 |
| SARIMA arima() | (3, 1, 2) | (0, 1, 1) | 7 | 2910.30 / 2946.99 | 0.46 | -1448.15 | 0.49 |
| SARIMA arima() | (3, 1, 2) | (1, 1 ,1) | 7 | 2905.94 / 2947.87 | 0.46 | -1444.97 | 0.79 |
| **SARIMA arima()** | **(2, 1, 2)** | **(1, 1 ,1)** | **7** | **2904.69 / 2941.38** | **0.46** | **-1445.35** | **0.68** |
| SARIMAX arima() [with step] | (2, 1, 2) | (1, 1, 1) | 7 | 2904.90 / 2946.83 | 0.46 | -1444.45 | 0.69 |
| **SARIMAX arima() [with step]** | **(1, 1, 1)** | **(1, 1, 1)** | **7** | **2907.80 / 2939.18** | **0.46** | **-1447.86** | **0.49** |
| SARIMAX arima() [with step and ramp] | (2, 1, 2) | (1, 1, 1) | 7 | 2906.53 / 2953.70 | 0.46 | -1444.26 | 0.68 |
